# Interpretable ECG manifolds link longitudinal electrical phenotypes to heart-failure risk in hypertrophic cardiomyopathy

**DOI:** 10.64898/2026.11.02.26367092

**Authors:** Pietro Cerveri, Marion Taconnè, Valentina Corino, Luca Mainardi

## Abstract

Hypertrophic cardiomyopathy (HCM) produces heterogeneous electrical phenotypes whose continuous organization, longitudinal evolution and clinical relevance remain poorly defined. We developed complementary variational-autoencoder representations to map HCM ECG morphology relative to physiological variation and characterize within-subject change. Using 5,194 normal-reference ECGs and 2,594 longitudinal visits from 650 individuals with HCM, we found that HCM overlapped the physiological manifold but extended towards its periphery, while an HCM-oriented manifold largely reorganized shared electrical modes. Mixed-effects models identified longitudinal drift in selected phenotypic axes. A lateral/frontal QRS-force coordinate remained time-dependent after censoring septal reduction therapy, atrial fibrillation and pacing, and lower values were associated with subsequent heart-failure risk in baseline and time-updated analyses. A time-conditioned change VAE identified four interpretable within-subject modes aligned with longer-term trajectories. Sarcomeric genotype was associated with phenotype position but not trajectory rate.

## 1. Introduction

Understanding how disease expression evolves over time is a central challenge in hypertrophic cardiomyopathy (HCM). The condition encompasses heterogeneous genetic substrates, electrical phenotypes and clinical trajectories [6, 1, 21], with some individuals remaining stable for decades, whereas others developing heart failure or life-threatening arrhythmias. Although pathogenic sarcomeric variants contribute to this heterogeneity, they do not fully determine an individual’s disease course [20, 22]. Relating phenotypic variation to longitudinal evolution therefore requires representations that capture both a patient’s current electrical state and its trajectory over time.

The 12-lead electrocardiogram (ECG) provides an accessible, repeatable measurement of cardiac electrical expression. In HCM, depolarization and repolarization abnormalities relate to structural phenotype and myocardial fibrosis and can precede overt hypertrophy in sarcomeric variant carriers [8, 12]. Computational methods have extended this information beyond conventional measurements, enabling HCM detection, identification of electrical subgroups and extraction of interpretable waveform representations [18, 15, 17]. Supervised ECG models have also tracked treatment-related changes and longitudinal variation in echocardiographic risk features [14, 7, 26, 28, 24, 5]. While these approaches demonstrate the rich clinical information contained in serial ECGs, having been optimized for predefined targets, they do not directly describe the multiple morphological directions along which electrical phenotypes evolve.

To address this limitation, we learn the geometry of ECG variation directly from waveform morphology using variational autoencoders (VAEs), without supervision from genotype, clinical severity or outcomes. In a learned manifold, a low-dimensional representation organizing waveform variation, each recording defines an electrical phenotype, and successive recordings trace its trajectory. This formulation allows us to ask whether structure learned from ECG morphology contains information about subsequent clinical course that was not supplied during training, while distinguishing departure from normal electrical variation, heterogeneity within HCM, and change within an individual. We investigate this possibility using base-line and serial ECGs from the Sarcomeric Human Cardiomyopathy Registry (SHaRe) [11], together with an independent reference set of normal ECGs from PTB-XL [27] and an external HCM cohort from MIMIC-IV-ECG [9]. Variational autoencoders learn separate manifolds of normal ECG morphology and of variation within HCM, while MIMIC-IV ECGs are projected into the frozen representations to assess external reproducibility. Serial recordings are projected into these fixed latent spaces to characterize longitudinal trajectories. A complementary time-conditioned model learns waveform changes between consecutive visits while accounting for elapsed time. Genotype, clinical events and outcomes are introduced exclusively after representation learning, enabling their relationships with the resulting electrical phenotypes to be assessed without defining those phenotypes in advance (Fig. 1). Our aim is to determine whether unsupervised ECG representations can characterize HCM electrical phenotype across complementary levels, including deviation from physiological variation, heterogeneity within established HCM and longitudinal within-patient change. We further examine whether these representations identify interpretable and reproducible electrical trajectories, how these trajectories relate to major clinical transitions and sarcomeric genotype, and whether selected phenotypes are associated with subsequent clinical outcomes. More broadly, we ask whether serial ECGs can provide a state-and-change representation of HCM without optimizing the representation for predefined diagnostic or prognostic targets.

**Figure 1:**
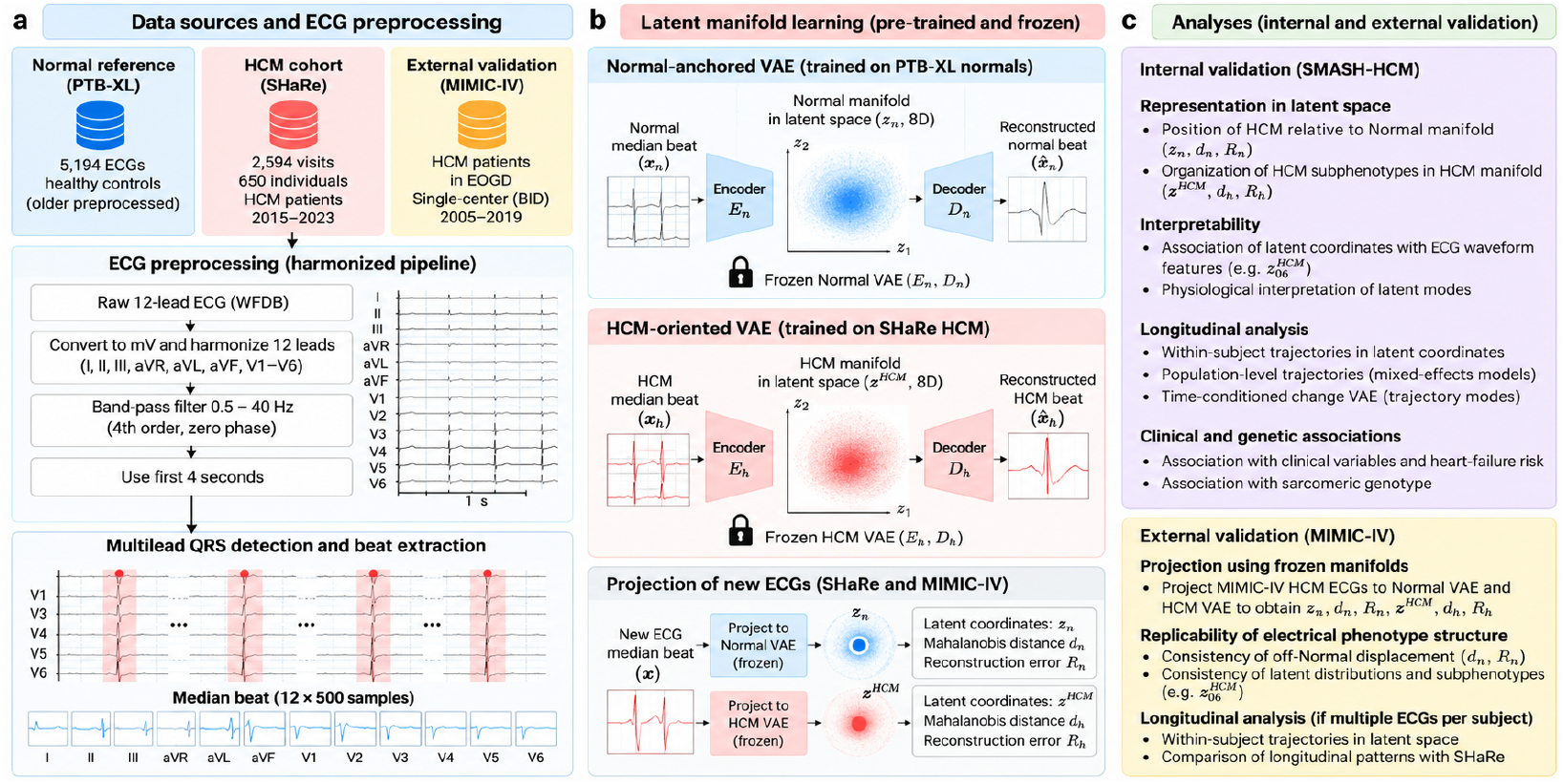
Overview of the study framework and external validation. **a**, Data sources and harmonized ECG preprocessing for Normal-reference and HCM cohorts. **b**, Training of complementary Normal-anchored and HCM-oriented VAEs, followed by projection of HCM ECGs to obtain latent coordinates, manifold distances and reconstruction-error measures; serial ECGs were additionally analysed through frozen-manifold trajectories and a time-conditioned Delta-VAE. **c**, Post hoc physiological, longitudinal, genetic and clinical analyses in SHaRe, together with external evaluation of the frozen representations in MIMIC-IV. No diagnostic classifier was trained or evaluated.

## 2. Methods

### 2.1. Study design and HCM cohort harmonization

The HCM cohort was derived from the Sarcomeric Human Cardiomyopathy Registry (SHaRe). Methods regarding SHaRe data collection have been previously described [11]. The HCM dataset initially contained 2,705 raw ECG identifiers. Identity reconciliation and removal of 107 redundant ECG identifiers yielded 2,598 unique visits from 650 coherent subjects. Of these, 649 subjects contributed an eligible baseline ECG for median-beat morphological analysis, with one baseline record excluded because it did not meet the prespecified minimum requirement of at least two usable aligned beats. Longitudinal visits were chronologically ordered within each coherent analysis subject. Four visits failed the minimum quality requirement for latent projection, leaving 2,594 valid visits. The primary longitudinal cohort required at least two valid visits at distinct time points and contained 493 subjects and 2,437 visits; the three-or-more-visit sensitivity cohort contained 376 subjects and 2,203 visits. Clinical characterization was restricted to the 623 subjects with an exact non-ambiguous ECG-to-SHaRe mapping. Base-line was defined by the first valid waveform ECG. Clinical-history variables were referenced to this baseline when event timing was available, and echocar-diographic measurements were obtained from the examination nearest to the baseline ECG within a ±7-day window.

### 2.2. Normal-reference cohort

The physiological reference cohort was derived from PTB-XL [27]. Eligible ECGs were required to have the SCP-ECG code `NORM` with likelihood 100, with no additional diagnostic SCP codes other than the physiological rhythm codes `SR`, `SBRAD`, `STACH` or `SARRH`. Subjects were additionally restricted to 18– 90 years of age, and only ECGs marked as human-validated in PTB-XL were retained. These criteria identified 5,417 eligible ECG records from 5,194 unique subjects. To avoid repeated observations from the same individual in the physiological reference model, one ECG was selected per subject, using the earliest available recording when multiple eligible records were present, yielding 5,194 Normal-reference ECGs. The Normal cohort was used to train and evaluate the Normal-anchored VAE and was not used to train either the HCM-oriented static VAE or the longitudinal Delta-VAE.

### 2.3. ECG harmonization and median-beat construction

ECGs were analysed using a 4-s segment corresponding to the first 2,000 samples of each record, featuring a sampling rate of 500 Hz. Each lead was filtered using a fourth-order Butterworth band-pass filter from 0.5 to 40 Hz with zero-phase application. Common multilead QRS fiducials were detected across the 12 leads. Beat windows extended from −350 ms to +650 ms relative to each detected multilead QRS fiducial, producing 500 samples per lead. A minimum of two usable beats was required for longitudinal latent projection. Median-beat morphology was analysed separately from rhythm and beat-to-beat descriptors. Consequently, latent morphology coordinates should not be interpreted as intrinsic heart-rate or rhythm representations even when empirical associations with timing-related features are observed.

### 2.4. Variational autoencoder framework

All ECG representation models were based on variational autoencoders (VAEs), in which an encoder maps each input waveform *X* to the parameters of an approximate Gaussian posterior distribution in latent space, *q*_*ϕ*_(*z*|*X*) = *N* (*µ*_*ϕ*_(*X*), 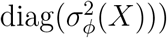. During training, latent samples were obtained using the reparameterization trick, *z* = *µ*_*ϕ*_(*X*) + *σ*_*ϕ*_(*X*) ⊙ *ϵ*, with *ϵ* ∼ *N* (0, *I*), and a decoder reconstructed the input waveform from the sampled latent representation. Models were optimized using a *β*-VAE objective combining mean-squared reconstruction error with Kullback–Leibler regularization towards a unit Gaussian prior,

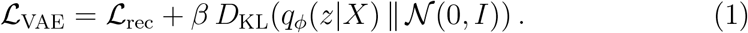

The reconstruction term promotes preservation of ECG morphology, whereas the KL term regularizes the latent representation and encourages a continuous structured latent space. For deterministic downstream analyses, latent coordinates were represented by the posterior mean *µ*_*ϕ*_(*X*) rather than by stochastic latent samples. Latent dimensions were considered active when the variance of their posterior means in the corresponding training set exceeded 0.01. Effective latent dimensionality was quantified from the eigenvalues *λ*_*i*_ of the training-set posterior-mean covariance matrix using the participation ratio, 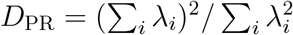.

### 2.5. Normal-anchored β-VAE

The encoder of the Normal-anchored *β*-VAE consisted of four one-dimensional convolutional blocks with channel progression 12–32–64–96–128 and temporal downsampling from 500 to 31 samples, followed by a 96-unit fully connected layer producing the posterior mean and log-variance; the decoder mirrored this architecture using transposed convolutions. Hidden convolutional blocks used GroupNorm and SiLU activations. The VAE was trained exclusively on Normal-reference median beats. Median-beat amplitudes were standardized using a single global mean and standard deviation estimated exclusively from the Normal training set. The same transformation was sub-sequently applied to Normal validation and test ECGs and to all HCM ECGs. No per-record or per-lead amplitude normalization was applied, thereby pre-serving relative voltage information and cross-lead amplitude relationships. The input shape was 12×500, the latent dimensionality was 16 and the model contained 939,116 trainable parameters. A *β*-VAE objective was used, with *β* linearly increased during the first 30 epochs to a target value of 0.01. Model selection based on the Normal validation set began once the target *β* was reached. HCM ECGs were projected after training with all model weights frozen. The latent posterior mean was used as the deterministic representation for downstream analyses. A Mahalanobis distance *d*_*N*_ was computed relative to the Normal-training posterior-mean distribution using a Ledoit–Wolf covariance estimate. Physical reconstruction mean squared error was calculated after inversion of the training-set scaling transformation. Because Normal and HCM data originated from different source datasets, HCM-versus-Normal reconstruction differences and any descriptive discrimination metrics were not interpreted as diagnostic validation.

### 2.6. HCM-oriented static β-VAE

The encoder of the HCM-oriented static *β*VAE consisted of four one-dimensional convolutional blocks with channel progression 12–16–32–48–64 and temporal downsampling from 500 to 31 samples, followed by a 48-unit fully connected layer producing the posterior mean and log-variance; the decoder mirrored this architecture using transposed convolutions. Hidden convolutional blocks used GroupNorm and SiLU activations. The model was trained on one baseline median-beat ECG per eligible HCM subject. Median-beat amplitudes were standardized using a single global mean and standard deviation estimated exclusively from the HCM training set and applied un-changed to the validation and test sets. No per-record or per-lead amplitude normalization was performed. Subject-wise HCM train, validation and test partitions contained 454, 97 and 98 ECGs, respectively. Normal-reference ECGs were not used for training or model selection. The input shape was 12 × 500, the latent dimensionality was 8 and the model contained 236,988 trainable parameters. A *β*-VAE objective was used, with *β* linearly increased during the first 30 epochs to a target value of 0.01. Model selection based on the HCM validation set began once the target *β* was reached. The purpose of this model was to characterize continuous electrical heterogeneity within HCM rather than to discriminate HCM from Normal ECGs. A Mahalanobis distance *d*_HCM_ was computed relative to the HCM-training posterior-mean distribution. Physiological interpretation of HCM latent axes used decoder traversals and associations with ECG morphology features extracted from real median beats.

### 2.7. Cross-manifold integration

To determine whether HCM-oriented coordinates represented morphology independent of the Normal-anchored space, each frozen HCM latent coordinate was predicted from the 16-dimensional Normal-anchored latent vector using ridge regression with held-out evaluation. High cross-manifold predictability was interpreted as evidence of HCM-oriented reparameterization or recombination of electrical modes present in the physiological reference space. As a descriptive measure of relative reconstruction burden, we additionally calculated Δ*RE* = *RE*_*N*_ − *RE*_HCM_, where *RE*_*N*_ and *RE*_HCM_ denote physical reconstruction errors from the frozen Normal-anchored and HCM-oriented VAEs, respectively. Associations between HCM latent coordinates and Δ*RE* were used to identify phenotypes linked to morphology that was relatively less well represented by the Normal-anchored model. Because the two autoencoders differed in training population and model capacity, Δ*RE* was interpreted descriptively and not as a calibrated measure of disease specificity. Residual components were therefore treated as off-Normal or not-explained-by-Normal-representation components and were not labelled as disease-specific without additional evidence.

### 2.8. Longitudinal projection through frozen static manifolds

All valid HCM longitudinal median beats were projected through both frozen static VAEs without additional representation learning. For each visit we retained the Normal-anchored and HCM-oriented posterior-mean latent coordinates, *d*_*N*_, *d*_HCM_, physical reconstruction errors and selected region-specific residual metrics. Baseline projection consistency was checked against the static baseline analyses before longitudinal modelling.

### 2.9. Mixed-effects analysis of longitudinal electrical trajectories

For each prespecified primary outcome, we fitted a linear mixed-effects model

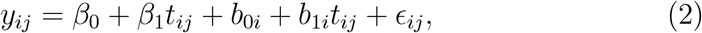

where *t*_*ij*_ was time in years from the subject’s first valid ECG, *β*_1_ was the population-average annual change and *b*_1*i*_ was the subject-specific deviation from that slope. Models were fitted by restricted maximum like-lihood with random intercept and random slope; a random-intercept-only fallback was allowed only when the random-slope model failed and was explicitly flagged. The 28 primary variables comprised 16 Normal-anchored latent coordinates, eight HCM-oriented latent coordinates, *d*_*N*_, *d*_HCM_ and the two model-specific physical reconstruction errors. Benjamini–Hochberg false-discovery-rate (FDR) correction was applied jointly across these 28 primary time-slope tests. Patient-specific BLUP slopes were exported for successfully fitted random-slope models. Independent subject-wise ordinary-least-squares (OLS) slopes and first-to-last annualized changes were calculated as model-independent descriptive complements. A sensitivity analysis repeated the primary mixed-effects analysis in subjects with at least three visits. To assess whether longitudinal electrical trajectories were sensitive to major clinical transitions capable of modifying ECG morphology, we performed event-censoring sensitivity analyses using clinical data from the SHaRe subset available to the present study through the SMASH-HCM project. Analyses were restricted to subjects with an exact ECG-to-SHaRe mapping. Septal reduction therapy (SRT) was defined as surgical myectomy or alcohol septal ablation. Separate sensitivity cohorts were constructed by censoring ECG observations occurring at or after the recorded onset of SRT, atrial fibrillation (AF) or pacemaker implantation, and by jointly censoring at the earliest of these three events. Subjects with an event recorded but unavailable event timing were excluded from the corresponding analysis, and only subjects retaining at least two valid ECGs at distinct time points after censoring were analysed. For each sensitivity cohort, the original longitudinal mixed-effects models were refitted without retraining either latent representation, using time from the first valid ECG as the fixed effect and subject-specific random intercepts and slopes. The selected trajectory-sensitive outcomes were 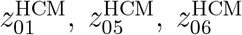 and distance from the Normal manifold (*d*_*N*_). Benjamini–Hochberg correction was applied across the four variables within each sensitivity analysis.

### 2.10. Clinical-risk association of the longitudinally robust 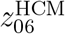 phenotype

To investigate the clinical relevance of the HCM-oriented coordinate showing the most robust longitudinal trajectory, we performed secondary time-to-event analyses focused on 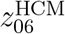. Analyses were restricted to subjects with an exact ECG-to-SHaRe mapping, a valid baseline 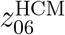 value and sufficient timing information to establish whether each clinical event occurred before or after the first valid ECG. The first valid ECG defined baseline and time zero, and subjects with the event of interest occurring before or at baseline were excluded from the corresponding risk set. Follow-up ended at the first occurrence of the endpoint or at the last available clinical follow-up in subjects without the event. The principal endpoint of this secondary clinical-risk analysis was the SHaRe heart-failure composite, defined as the first occurrence of left ventricular ejection fraction *<* 35%, New York Heart Association functional class III/IV symptoms, cardiac transplantation or left ventricular assist-device implantation. Baseline analyses additionally evaluated atrial fibrillation, unplanned cardiovascular hospitalization and the overall composite endpoint, with Benjamini–Hochberg false-discovery-rate correction applied across these four clinical outcomes. The ventricular-arrhythmic composite, defined as sudden cardiac death, resuscitated cardiac arrest or appropriate implantable cardioverter-defibrillator therapy, was analysed separately as an exploratory endpoint because of the limited number of incident events. For baseline analyses, 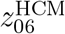 was standardized using the standard deviation of baseline 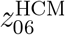 in the full exact-mapped cohort, and Cox proportional-hazards models were fitted both unadjusted and adjusted for baseline age and sex. Hazard ratios therefore represent the relative change in event hazard per 1-SD increase in 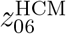. To determine whether serially available phenotype values retained prognostic information, we additionally fitted time-updated Cox models using a start–stop formulation in which the most recent observed 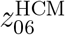 value was carried forward from each ECG until the subsequent ECG, event or censoring time. Only ECG information available before each event contributed to the corresponding risk interval. In a 30-day lag sensitivity analysis, each ECG-derived phenotype value became active 30 days after its acquisition, with follow-up correspondingly left-truncated at 30 days. This prevented phenotype measurements obtained within 30 days before an event from contributing to the corresponding risk interval. Robustness of the time-updated estimates was further assessed using 2,000 subject-level bootstrap resamples. To explore residual risk after septal reduction therapy (SRT), landmark analyses were performed using the first eligible ECG obtained 30– 180 days after SRT, with a secondary 30–365-day window. Only subjects free of the endpoint at the landmark entered the corresponding risk set, and subsequent events were counted from the landmark ECG. These post-SRT analyses were considered exploratory because of the small number of eligible subjects and subsequent events.

### 2.11. HCM-only longitudinal Delta-VAE

The longitudinal change VAE was trained exclusively on consecutive pairs of valid HCM visits. For each pair,

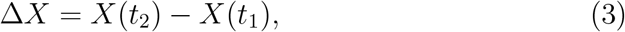

where *X*(*t*) was the 12-lead median beat in physical mV units. The primary model encoded the signed waveform difference Δ*X* directly rather than the interval-normalized quantity Δ*X/*Δ*t*, thereby avoiding amplification of measurement and alignment noise at short intervals. Elapsed inter-visit time *τ* entered explicitly as a conditioning variable as:

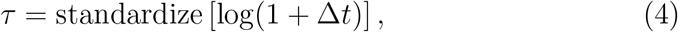

where the mean and standard deviation used for standardization were estimated exclusively from the training partition and subsequently applied unchanged to validation and test pairs. Signed waveform differences were similarly standardized using a single global mean and standard deviation estimated from training pairs only. The conditional encoder comprised four one-dimensional convolutional blocks with channel progression 12–16–32–48– 64 and temporal downsampling from 500 to 31 samples, followed by fully connected layers of 64 and 48 units. The standardized time-conditioning variable *τ* was concatenated to the encoded representation before estimation of the posterior mean and log-variance and was concatenated again to the latent vector before decoding. The decoder mirrored the convolutional hierarchy using transposed convolutions with GroupNorm and SiLU activations in its hidden blocks. The resulting conditional representation was:

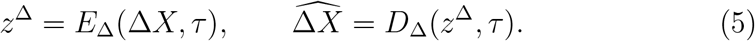

Subjects, rather than ECG pairs, were assigned to train, validation and test partitions to prevent information leakage across repeated observations from the same individual. During training, each pair was weighted inversely to the number of training pairs contributed by its subject, with weights rescaled to unit mean, thereby limiting disproportionate influence from subjects with more repeated ECGs. The model used 1,916 consecutive pairs from 491 subjects, a four-dimensional latent representation and 271,396 trainable parameters. The latent dimensionality was set to four to provide a compact representation of within-subject ECG change while retaining sufficient capacity to capture multiple distinct morphological change modes. A *β*-VAE objective was used, with *β* linearly increased during the first 30 epochs to a target value of 0.01; model selection based on the validation loss began once the target *β* was reached. All four latent dimensions remained active after training. The longitudinal latent variables were interpreted as model-dependent modes of ECG morphological change rather than as clinical progression scores.

### 2.12. Physiological interpretation of longitudinal change modes

Classical ECG features were not extracted directly from Δ*X* because a difference waveform is not itself a physiological ECG. Instead, features were extracted independently from *X*(*t*_1_) and *X*(*t*_2_) and transformed into observed feature changes Δ*f* = *f* (*t*_2_) − *f* (*t*_1_). Spearman associations between each Delta-VAE posterior-mean coordinate and each observed feature change were calculated separately in the training, validation and held-out test subsets. Benjamini–Hochberg false-discovery-rate correction was applied globally across all latent–feature associations within each subset, with the held-out test associations used as the primary basis for physiological interpretation. Decoder-based interpretation was performed by varying one latent coordinate from −3 to +3 empirical training-set standard deviations while fixing all remaining latent coordinates at zero. The inter-visit interval was fixed at the median Δ*t* of the training pairs. Decoded waveform changes were added to real HCM reference beats drawn from the training set, and ECG features were remeasured on the resulting synthetic beats. For each traversal level, feature changes were quantified relative to the corresponding zero-latent decoded trajectory at the same Δ*t* and summarized across reference beats. A latent–feature relationship was considered a strong internal interpretation when the held-out test association had |*ρ*| ≥ 0.20 and global FDR *q <* 0.05, the decoder traversal showed *R*^2^ ≥ 0.80, and the directions of the observed and traversal-based effects were concordant.

### 2.13. Cluster-aware robustness of physiological anchors

Because some subjects contributed multiple consecutive ECG pairs, four physiological anchor associations identified in the preceding interpretation analysis, one for each Delta-VAE latent mode, were further assessed using analyses that accounted for within-subject clustering. All analyses were restricted to the held-out Delta-VAE test subset. Pair-level Spearman correlations were retained as descriptive effect estimates, whereas inference was based on three complementary subject-aware procedures: 10,000 subject-cluster bootstrap resamples retaining all pairs from each resampled subject; subject-level aggregation using the median latent coordinate and median feature change for each subject, with 20,000 permutation tests and Benjamini– Hochberg correction across the four selected anchor associations; and 10,000 Monte Carlo repetitions in which one pair was randomly selected per subject. An anchor was considered robust when the subject-cluster bootstrap 95% confidence interval excluded zero, the subject-level association preserved the expected direction with FDR *q <* 0.05, and at least 90% of one-pair-persubject repetitions preserved the expected sign. Because these four anchors were selected from the preceding physiological interpretation analysis, these analyses were considered cluster-aware robustness assessments rather than statistically independent validation.

### 2.14. Interval-normalized rate sensitivity analysis

As a secondary sensitivity analysis, a separately trained four-dimensional VAE was fitted to interval-normalized waveform change Δ*X/*Δ*t*. To reduce amplification of measurement and alignment noise at short inter-visit intervals, pairs separated by less than 0.25 years were excluded. The rate model was not conditioned on Δ*t* and therefore represented annualized wave-form change directly rather than learning an explicit dependence on elapsed time. The resulting analysis included 1,775 consecutive ECG pairs from 488 HCM subjects. Physiological interpretation used annualized feature changes Δ*f/*Δ*t* and the same held-out association and decoder-traversal framework used for the primary Delta-VAE. Decoder traversals were interpreted on a one-year-equivalent scale. Because independently trained VAE coordinates may permute, invert or rotate, the sensitivity analysis was evaluated for recovery of the same broad physiological change domains rather than for one-to-one correspondence of latent indices.

#### Integration of Delta-VAE change modes with patient-specific trajectories

For each subject, the signed median of each Delta-VAE coordinate across all consecutive ECG pairs was used as a subject-level summary of typical change-mode expression. These four summaries were compared with patient-specific BLUP slopes for the selected longitudinal targets 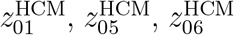 and *d*_*N*_, yielding 16 subject-level associations. Spearman correlation was used, subject-level bootstrap 95% confidence intervals were calculated, and Benjamini–Hochberg FDR correction was applied across the 16 associations. The primary integration analysis included 491 subjects. Two sensitivity analyses were performed. First, the same associations were evaluated in the 73 subjects belonging to the held-out Delta-VAE test split to assess whether the observed alignment extended beyond subjects used for model training. Second, BLUP slopes were replaced by empirical subject-wise OLS slopes among 376 subjects with at least three valid ECG visits, providing a model-independent sensitivity analysis to BLUP shrinkage. These analyses quantify alignment between complementary longitudinal representations and are not considered statistically independent validation because the Delta-VAE summaries and trajectory slopes are derived from overlapping serial ECG observations.

### 2.15. Genetic analysis

The primary sarcomeric genotype definition classified subjects as Sarc+ when an exact non-ambiguous HCM mapping contained at least one pathogenic or likely pathogenic variant in a prespecified core sarcomeric gene (*ACTC1, MYBPC3, MYH7, MYL2, MYL3, TNNI3, TNNT2* or *TPM1*). Sarc− subjects required an exact non-ambiguous negative genetic result and absence of pathogenic or likely pathogenic variants. Subjects with ambiguous mapping, VUS-only findings or other non-primary genotype categories were excluded from the primary contrast. The primary longitudinal genetic cohort included 320 subjects and 1,720 visits. For 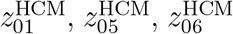 and *d*_*N*_, we fitted linear mixed-effects models of the form

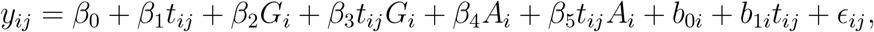

where *t*_*ij*_ was time in years from the first valid ECG, *G*_*i*_ was coded as 0 for Sarc− and 1 for Sarc+, and *A*_*i*_ was baseline age centered at the cohort mean. Subject-specific random intercepts and time slopes were included. Under this parameterization, *β*_1_ represents the annual trajectory in Sarc− subjects, *β*_3_ represents the difference in annual trajectory between Sarc+ and Sarc− subjects, and *β*_1_ + *β*_3_ represents the annual trajectory in Sarc+ subjects. The genotype-by-time interaction *β*_3_ was the primary estimand, with Benjamini–Hochberg FDR correction applied across the four prespecified interaction tests. The genotype main effect *β*_2_ represents the Sarc+ versus Sarc− difference at baseline at the cohort mean baseline age and was considered secondary.

### 2.16. External evaluation in MIMIC-IV

An independent HCM cohort was derived from MIMIC-IV and processed using the same median-beat representation in the primary SHaRe analysis. ECGs were harmonized to 500 Hz, band-pass filtered at 0.5–40 Hz using a fourth-order zero-phase Butterworth filter, and represented as QRS-aligned 1-s median beats. Probable duplicate acquisitions were conservatively defined as within-subject ECG pairs with normalized RMSE ≤ 0.10, Pearson correlation ≥ 0.995 and acquisition-time separation ≤ 1 h, and were collapsed using a direct anchor-based rule without transitive chaining. After removal of 12 probable duplicate acquisitions, 5,694 ECGs from 335 subjects remained, of which 5,564 satisfied the prespecified requirement of at least two usable aligned beats. Eligible MIMIC median beats were projected through the frozen Normal-anchored and HCM-oriented VAEs using the original model weights, training-derived scaling parameters and latent reference distributions; MIMIC data were not used for retraining, fine-tuning, rescaling or covariance estimation. Baseline manifold transport was evaluated using the first valid ECG from each subject, with empirical 95% coverage thresholds derived exclusively from the corresponding original training distributions. Distributional transport of the prespecified 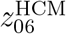 phenotype was assessed using standardized mean differences with subject-level bootstrap confidence intervals and Kolmogorov–Smirnov statistics. For longitudinal external replication, temporally clustered ECGs were consolidated into bounded 30-day windows anchored to the first ECG in each window, without transitive chaining, and one representative ECG was retained per window. Subjects with at least two representative ECGs at distinct time points were included. The pre-specified primary analysis used a linear mixed-effects model of 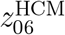 versus time from the first valid ECG, with subject-specific random intercepts and slopes and restricted maximum-likelihood estimation. Sensitivity analyses used 7- and 90-day windows, required at least three visits or at least one year of follow-up, and repeated the model without clinical-window consolidation. Robustness to unequal ECG sampling density was additionally assessed using one empirical OLS slope per subject, subject-level bootstrap confidence intervals for the median slope, exact sign tests and Wilcoxon signed-rank tests.

### 2.17. Statistical analysis

Continuous associations used Spearman rank correlation unless otherwise specified. Multiple testing was controlled with the Benjamini–Hochberg FDR procedure within prespecified analysis families. Mixed-effects models used restricted maximum likelihood unless otherwise stated. Patient-specific BLUP slopes are shrinkage estimates and were not interpreted as independent raw slopes. Bootstrap and permutation procedures used subject-level resampling or subject-level aggregation when required to account for repeated observations or repeated ECG pairs. For visualization only in Fig. 6b, extremely isolated BLUP slopes with an absolute robust MAD-based *z*-score *>* 8 were omitted from the scatter display to avoid compression of the remaining observations; all subjects were retained in the reported correlations, confidence intervals and multiple-testing procedures. All reported tests were two-sided unless explicitly specified otherwise. Analyses were performed using Python 3.12.10, PyTorch 2.13.0+cu132, NumPy 2.5.3, SciPy 1.18.1, pandas 3.0.6, statsmodels 0.15.0 and scikit-learn 1.9.1. The complete frozen analysis environment is provided with the reproducibility archive. For the secondary clinical-risk analyses, Cox proportional-hazards models were used as specified above; hazard ratios were expressed per 1-SD increase in 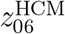, and bootstrap resampling was performed at the subject level.

## 3. Results

### 3.1. Study population and analysis framework

The HCM cohort was derived from the Sarcomeric Human Cardiomyopathy Registry (SHaRe) [11]. After identity harmonization and removal of redundant ECG identifiers, the dataset comprised 2,598 unique visits from 650 coherent subject (Extended Data Table 4). Of these, 649 subjects contributed an eligible baseline ECG for median-beat morphological analysis, with one baseline record excluded because it did not meet the prespecified minimum requirement of at least two usable aligned beats. Overall, 2,594 visits passed the longitudinal projection quality criteria; 493 subjects contributed at least two valid visits and 376 contributed at least three. Of the 649 waveform subjects in the SHaRe cohort, 623 had an exact non-ambiguous ECG-to-SHaRe mapping and constituted the clinical-characterization cohort. The strict Normal reference comprised 5,194 subjects selected from PTB-XL [27]. Static analyses used one baseline ECG per subject, whereas longitudinal analyses retained all eligible HCM visits. The analysis framework separated physiological reference modelling, within-HCM static phenotyping, longitudinal projection through frozen manifolds and direct modelling of within-subject waveform change, followed by clinical and genetic interrogation of the resulting electrical phenotypes. For external evaluation, an independent HCM cohort derived from MIMIC-IV [9] comprised 5,706 ECG records from 335 subjects, of which 5,576 satisfied the prespecified median-beat quality criterion and were retained for projection through the frozen Normal-anchored and HCM-oriented manifolds.

### 3.2. A Normal-anchored ECG manifold defines physiological morphological variation

The Normal-anchored VAE was trained exclusively on Normal-reference median beats and used a 16-dimensional latent representation. All 16 dimensions were active, with an effective participation-ratio dimension of approximately 14.5. Reconstruction error was substantially larger for HCM than for held-out Normal ECGs: the median physical reconstruction mean squared error was 0.00692 mV^2^ in HCM and 0.000801 mV^2^ in the Normal test set. These differences are interpreted as descriptive evidence that HCM morphology is less well represented by the physiological reference manifold, not as diagnostic performance because the HCM and Normal cohorts originate from different data sources. Latent traversal and feature association analyses linked individual Normal-anchored coordinates to interpretable electrical domains, including LV-oriented depolarization forces, ventricular activation geometry, coupled depolarization–repolarization morphology and frontal repolarization orientation (Extended Data Fig. 4. HCM ECGs were not confined to a separate remote cluster; instead, their latent distribution overlapped the Normal manifold while being shifted and more dispersed towards its tails and periphery (Fig. 2). Representative 12-lead median-beat morphologies from Normal-reference and HCM subjects are shown in Extended Data Fig. 4.

**Figure 2:**
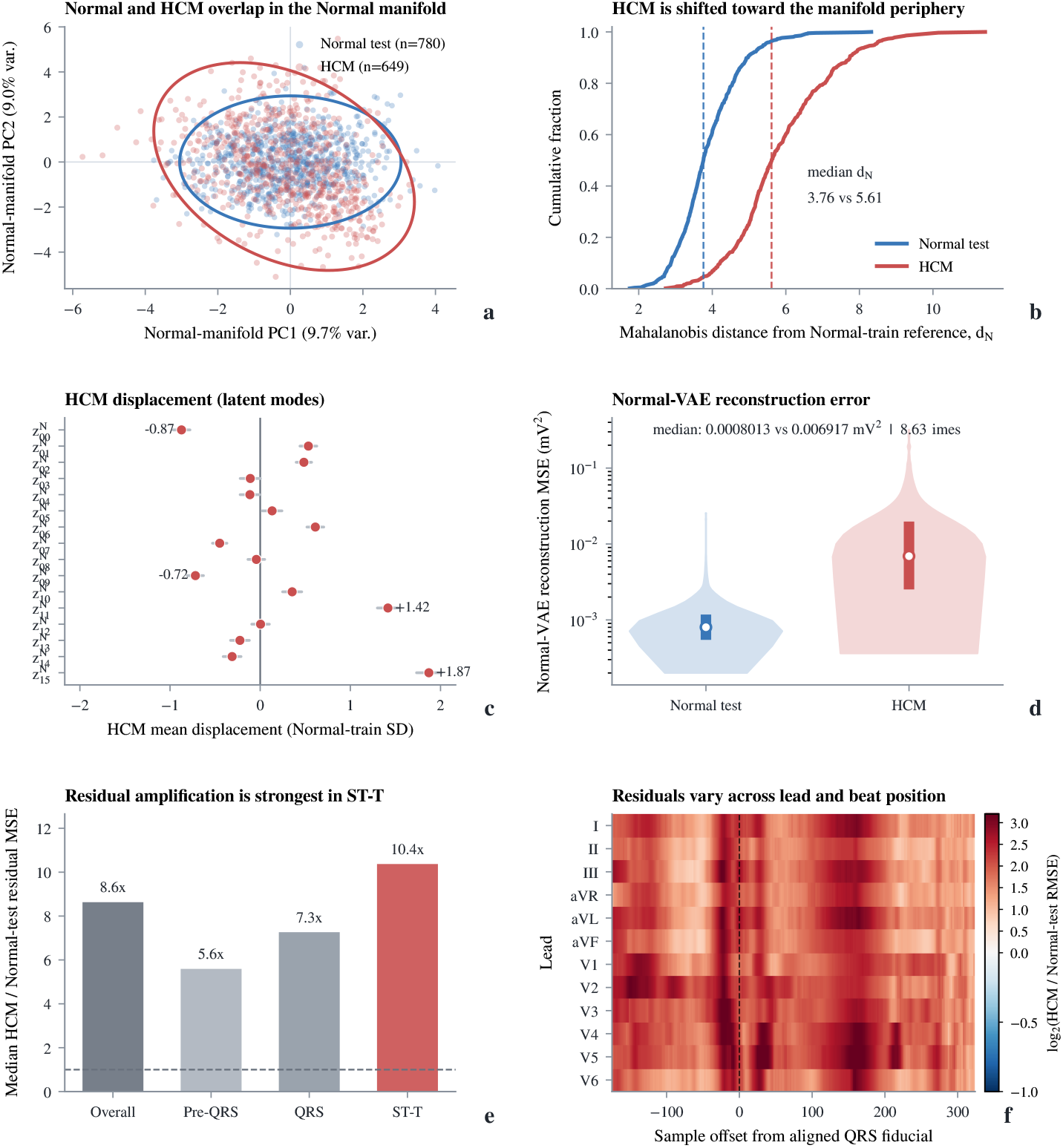
Normal-anchored representation of HCM ECG morphology. **a**, PCA projection of the Normal-anchored latent space showing held-out Normal and HCM ECGs. **b**, Distribution of the Mahalanobis distance from the Normal-training reference (*d*_*N*_), with HCM shifted toward more peripheral regions. **c**, Mean HCM displacement across the 16 Normal latent coordinates, expressed in Normal-training SD units. **d**, Reconstruction error of the frozen Normal-VAE in Normal-test and HCM ECGs. **e**, Regional amplification of reconstruction residuals in HCM across pre-QRS, QRS and ST–T segments. **f**, Lead- and beat-position-specific residual amplification, expressed as log_2_(HCM/Normal-test RMSE). HCM ECGs were used only after model freezing for projection and evaluation.

### 3.3. The HCM-oriented manifold reorganizes largely shared electrical variation

A separate eight-dimensional VAE was trained on baseline HCM median beats to characterize within-HCM electrical heterogeneity. All eight dimensions were active, with an effective participation-ratio dimension of 7.00. Physiological interpretation based on associations with ECG features measured in real HCM median beats and concordant decoder traversals identified continuous axes dominated by precordial QRS-force redistribution, precordial R/S transition balance, precordial repolarization, precordial depolarization force, global QRS magnitude, lateral/LV-oriented depolarization force, lateral/frontal QRS-force redistribution and mixed precordial repolarization–activation patterns (Extended Data Table 4)). Cross-manifold analysis showed that the HCM-oriented latent coordinates were substantially predictable from the Normal-anchored representation. Held-out ridge-regression *R*^2^ values ranged from 0.63 to 0.86 across the eight HCM latent axes, indicating that the HCM-oriented manifold is not well described as an orthogonal disease-only subspace. Instead, it predominantly reparameterizes and recombines electrical modes already represented within physiological ECG variation. Nevertheless, some HCM-oriented coordinates remained associated with morphology that was relatively poorly represented by the Normal-anchored model. In particular, the precordial repolarization coordinate 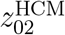 showed the strongest association with relative off-Normal reconstruction burden (*ρ* = −0.560 across all baseline HCM ECGs and *ρ* = −0.588 in the held-out HCM test set), despite being highly predictable from the Normal latent representation 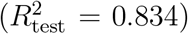. Thus, substantial cross-manifold predictability and relative off-Normal morphology are not mutually exclusive, but characterize complementary aspects of the HCM electrical phenotype (Fig. 3).

**Figure 3:**
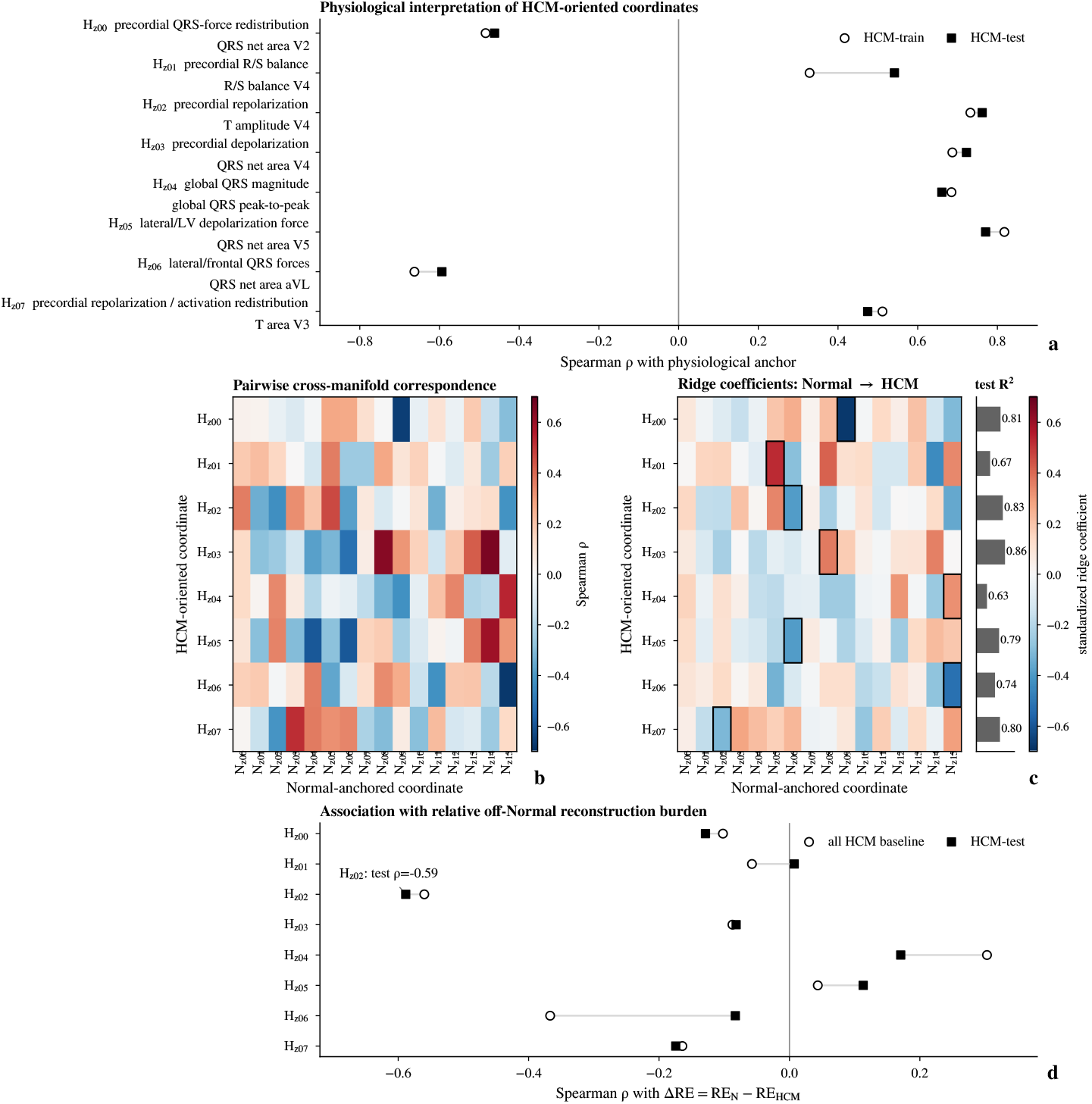
HCM-oriented latent phenotypes and their relationship to the Normal ECG manifold. **a**, Physiological interpretation of the eight HCM latent dimensions based on representative ECG-feature associations supported by concordant decoder traversals. **b**, Cross-manifold Spearman correlations between HCM and Normal latent coordinates. **c**, Multivariable ridge mapping from Normal to HCM latent space, with held-out *R*^2^. **d**, Association of HCM latent coordinates with relative off-Normal reconstruction burden (Δ*RE*).

### 3.4. HCM electrical phenotypes show longitudinal population-level trajectories

All valid longitudinal HCM ECGs were projected through the frozen Normal-anchored and HCM-oriented VAEs. Linear mixed-effects models with subject-specific random intercepts and slopes were fitted to 493 subjects with at least two valid ECGs, comprising 2,437 visits. Thirteen of 28 prespecified primary longitudinal outcomes were significant after false-discovery-rate correction. Within the HCM-oriented space, three coordinates showed significant population-average longitudinal change. The precordial R/S transition-balance coordinate 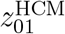 decreased by 0.0329 latent units per year (FDR *q* = 7.57 × 10^*−*5^), the lateral/LV-oriented depolarization-force coordinate 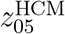 decreased by 0.0272 units per year (*q* = 9.16 × 10^*−*4^), and the lateral/frontal QRS-force coordinate 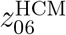 showed the strongest change, decreasing by 0.0531 units per year (*q* = 4.81 × 10^*−*12^). In contrast, the repolarization coordinate 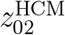 showed no detectable linear population trend. Distance from the Normal manifold (*d*_*N*_) increased by 0.0361 units per year (*q* = 0.00117), whereas distance within the HCM distribution remained broadly stable. Secondary residual analyses localized much of the longitudinal change to QRS/depolarization morphology rather than ST–T residual structure. A sensitivity analysis restricted to subjects with at least three visits preserved the direction of 26 of 28 primary slopes (Fig. 4). We next examined whether the principal longitudinal trajectories were sensitive to major clinical transitions capable of modifying ECG morphology. Of the 493 subjects in the primary longitudinal cohort, 475 had an exact ECG-to-SHaRe mapping, contributing 2,369 valid ECG visits; restricting the analysis to these subjects yielded estimates closely similar to the primary analysis. Censoring ECGs at septal reduction therapy (SRT) attenuated the annual slopes of 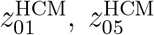 and *d*_*N*_, whereas 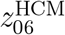 remained strongly associated with time (*β* = −0.0332 year^*−*1^, 95% CI −0.0464 to −0.0200, *q* = 3.3×10^*−*6^). Censoring at atrial fibrillation produced more modest attenuation, while censoring at pacemaker implantation had little effect. In the most conservative analysis, censoring at the earliest occurrence of SRT, atrial fibrillation or pacemaker implantation, 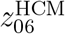 remained significantly time-dependent (*β* = −0.0311 year^*−*1^, 95% CI −0.0453 to −0.0168, *q* = 7.7 × 10^*−*5^), whereas the other three selected trajectories were attenuated and no longer significant after multiple-testing correction (Table 1). Together, these analyses indicate population-average longitudinal electrical evolution with substantial between-subject heterogeneity and differential sensitivity of individual electrical phenotypes to major clinical transitions. Longitudinal change along *z*^HCM^ was particularly robust to censoring of SRT, atrial fibrillation and pacing, whereas changes in 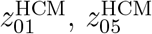 and *d*_*N*_ were partly attenuated. These population-level trajectories do not imply deterministic disease progression in individual patients.

**Figure 4:**
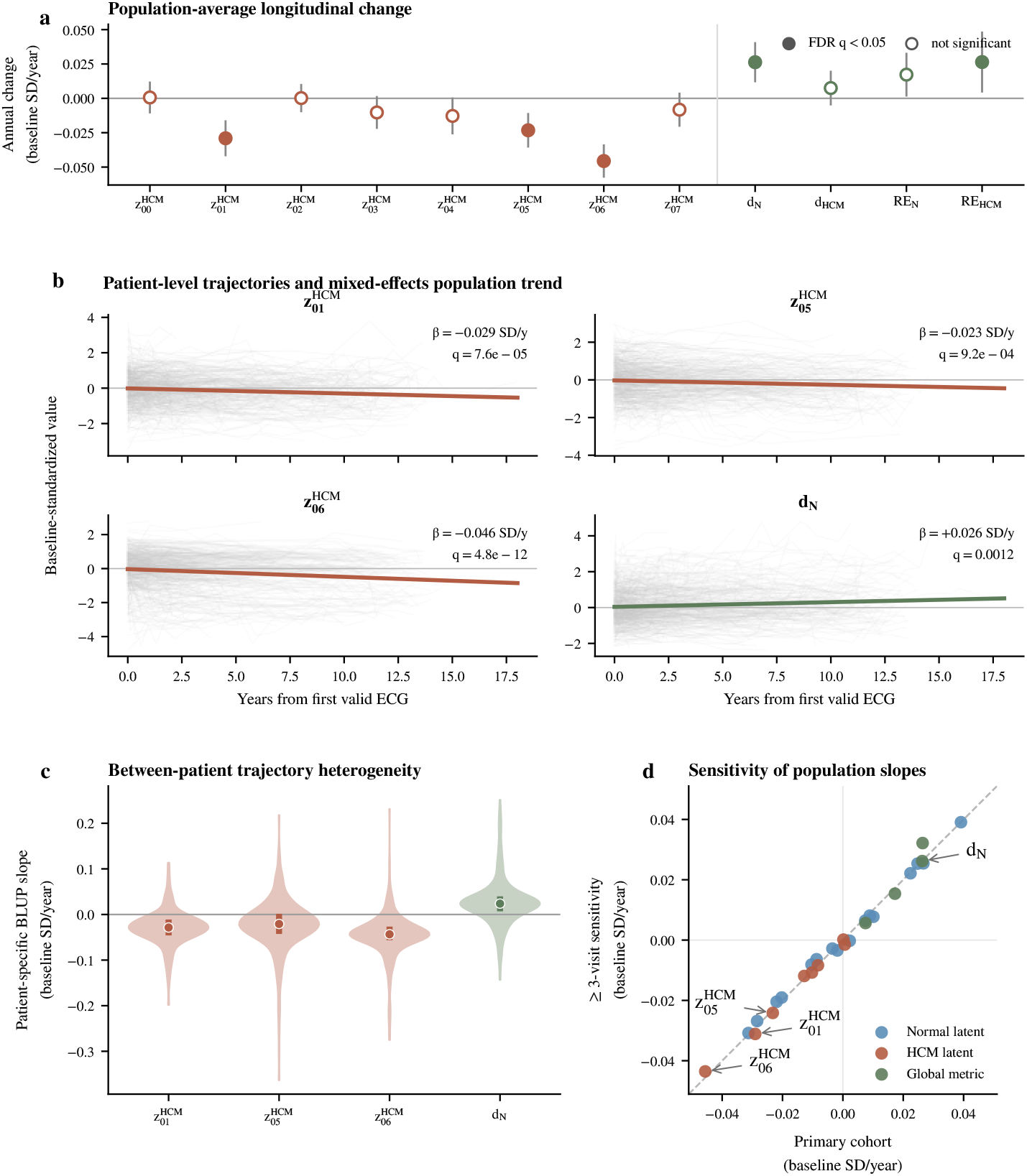
Population-level and patient-specific longitudinal electrical trajectories. **a**, Population-average annual changes in HCM-oriented latent coordinates and global manifold metrics, expressed in baseline standard-deviation units; filled markers indicate FDR *q <* 0.05. **b**, Individual longitudinal trajectories for 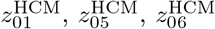 and distance from the Normal manifold (*d*_*N*_), with mixed-effects population trends overlaid. **c**, Distributions of patient-specific BLUP slopes for the same four outcomes, illustrating between-patient trajectory heterogeneity. **d**, Comparison of standardized population slopes between the primary cohort and the sensitivity analysis restricted to subjects with at least three valid visits.

**Table 1:** Sensitivity of longitudinal electrical trajectories to major clinical transitions. Linear mixed-effects models were refitted after restriction to subjects with exact ECG-to-SHaRe mapping and after censoring ECG observations at major clinical transitions. SRT denotes septal reduction therapy, defined as surgical myectomy or alcohol septal ablation. “Combined” denotes censoring at the earliest occurrence of SRT, atrial fibrillation or pacemaker implantation. Estimates are population-average annual slopes with 95% confidence intervals. Benjamini–Hochberg correction was applied across the four selected outcomes within each sensitivity analysis.

| Analysis | Outcome | Subjects / ECGs | Annual slope (95% CI) | FDR *q* |
|---|---|---|---|---|
| Reference | *z*<sub>01</sub><sup>HCM</sup> | 475 / 2,369 | −0.0328 (−0.0477 to −0.0180) | 2.96 × 10<sup>−5</sup> |
|  | *z*<sub>05</sub><sup>HCM</sup> |  | −0.0287 (−0.0437 to −0.0136) | 2.56 × 10<sup>−4</sup> |
|  | *z*<sub>06</sub><sup>HCM</sup> |  | −0.0525 (−0.0668 to −0.0382) | 2.79 × 10<sup>−12</sup> |
|  | *d*<sub>*N*</sub> |  | 0.0352 (0.0148 to 0.0555) | 7.07 × 10<sup>−4</sup> |
| SRT | *z*<sub>01</sub><sup>HCM</sup> | 444 / 2,013 | −0.0174 (−0.0332 to −0.0015) | 0.0544 |
|  | *z*<sub>05</sub><sup>HCM</sup> |  | −0.0156 (−0.0306 to −0.0007) | 0.0544 |
|  | *z*<sub>06</sub><sup>HCM</sup> |  | −0.0332 (−0.0464 to −0.0200) | 3.30 × 10<sup>−6</sup> |
|  | *d*<sub>*N*</sub> |  | 0.0095 (−0.0095 to 0.0286) | 0.326 |
| AF | *z*<sub>01</sub><sup>HCM</sup> | 410 / 1,937 | −0.0287 (−0.0445 to −0.0129) | 7.25 × 10<sup>−4</sup> |
|  | *z*<sub>05</sub><sup>HCM</sup> |  | −0.0220 (−0.0376 to −0.0065) | 0.00737 |
|  | *z*<sub>06</sub><sup>HCM</sup> |  | −0.0454 (−0.0601 to −0.0307) | 5.56 × 10<sup>−9</sup> |
|  | *d*<sub>*N*</sub> |  | 0.0199 (−0.0008 to 0.0405) | 0.0592 |
| Pacemaker | *z*<sub>01</sub><sup>HCM</sup> | 461 / 2,256 | −0.0302 (−0.0453 to −0.0151) | 1.79 × 10<sup>−4</sup> |
|  | *z*<sub>05</sub><sup>HCM</sup> |  | −0.0267 (−0.0418 to −0.0115) | 7.34 × 10<sup>−4</sup> |
|  | *z*<sub>06</sub><sup>HCM</sup> |  | −0.0520 (−0.0671 to −0.0369) | 5.49 × 10<sup>−11</sup> |
|  | *d*<sub>*N*</sub> |  | 0.0311 (0.0108 to 0.0515) | 0.00271 |
| Combined | *z*<sub>01</sub><sup>HCM</sup> | 379 / 1,656 | −0.0125 (−0.0290 to 0.0039) | 0.191 |
|  | *z*<sub>05</sub><sup>HCM</sup> |  | −0.0114 (−0.0266 to 0.0038) | 0.191 |
|  | *z*<sub>06</sub><sup>HCM</sup> |  | −0.0311 (−0.0453 to −0.0168) | 7.72 × 10<sup>−5</sup> |
|  | *d*<sub>*N*</sub> |  | 0.0021 (−0.0185 to 0.0227) | 0.839 |

### 3.5. 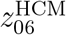 is associated with subsequent heart-failure risk

We next tested whether the HCM-oriented coordinate showing the most robust longitudinal trajectory was associated with subsequent clinical events. Among 574 subjects included in the baseline heart-failure risk analysis, 78 experienced a subsequent heart-failure event. Lower baseline 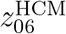 values were associated with higher heart-failure risk. After adjustment for baseline age and sex, each 1-SD increase in 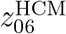 was associated with a 28% lower heart-failure hazard (HR 0.723, 95% CI 0.583–0.897, *p* = 0.0031, FDR *q* = 0.0125), corresponding to approximately 38% higher hazard per 1-SD decrease (Table 2). The association was preserved when 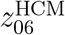 was treated as a time-updated exposure (574 subjects, 78 events; HR 0.743, 95% CI 0.609–0.908, *p* = 0.0037) and remained essentially unchanged after imposing a 30-day lag between ECG assessment and subsequent events (573 subjects, 77 events; HR 0.744, 95% CI 0.608–0.910, *p* = 0.0039). Subject-level bootstrap resampling (2,000 replicates) further supported the stability of the time-updated association, yielding a median HR of 0.745 (95% bootstrap CI 0.620–0.908); all bootstrap models converged without warnings, and the estimated 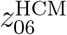 coefficient was negative in 99.7% of replicates. The association with ventricular-arrhythmic events was directionally consistent but based on substantially fewer events. Among 600 subjects included in the corresponding time-updated analysis, 16 experienced a subsequent ventricular-arrhythmic event; the age- and sex-adjusted HR was 0.580 per 1-SD increase in *z*^HCM^ (95% CI 0.406–0.828, *p* = 0.0027). This finding was therefore considered exploratory. Post-SRT landmark analyses were also underpowered: only 28 subjects had an eligible ECG 30–180 days after septal reduction therapy, yielding only three subsequent heart-failure events among 14 subjects remaining at risk. Extending the landmark window to 30–365 days increased the eligible cohort to 42 subjects, but only six heart-failure events occurred among 23 subjects at risk and no stable association with post-SRT 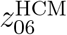 was observed. These data therefore precluded a reliable assessment of residual post-treatment risk. Collectively, these analyses identify 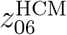 as a risk-associated electrical phenotype, with the strongest evidence for subsequent heart failure, but do not establish independent prognostic value or post-SRT residual-risk stratification.

**Table 2:**
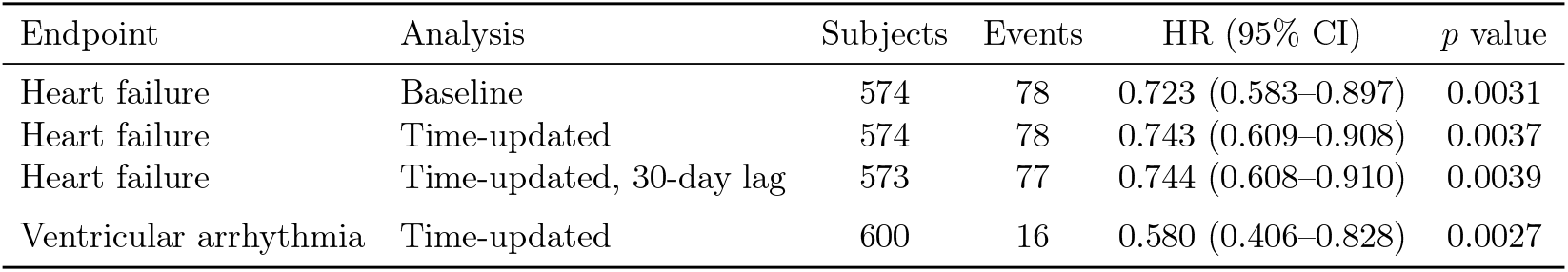
Association of 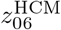 with subsequent clinical events. Hazard ratios were expressed per 1-SD increase in 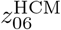 and adjusted for baseline age and sex. Heart failure was the principal endpoint reported in this secondary analysis. In the 30-day lag analysis, each ECG-derived phenotype value became active 30 days after acquisition, with follow-up left-truncated at 30 days. Ventricular-arrhythmic analyses were exploratory because of the limited number of incident events.

| Endpoint | Analysis | Subjects | Events | HR (95% CI) | *p* value |
|---|---|---|---|---|---|
| Heart failure | Baseline | 574 | 78 | 0.723 (0.583–0.897) | 0.0031 |
| Heart failure | Time-updated | 574 | 78 | 0.743 (0.609–0.908) | 0.0037 |
| Heart failure | Time-updated, 30-day lag | 573 | 77 | 0.744 (0.608–0.910) | 0.0039 |
| Ventricular arrhythmia | Time-updated | 600 | 16 | 0.580 (0.406–0.828) | 0.0027 |

### 3.6. An HCM-only longitudinal change manifold identifies four interpretable within-subject modes

To model within-subject change directly, we trained a four-dimensional conditional Delta-VAE based on the same VAE framework, applied to signed differences between consecutive HCM ECGs. The input was the signed, non-rate-normalized median-beat difference Δ*X* = *X*(*t*_2_) − *X*(*t*_1_), while the elapsed inter-visit interval Δ*t* entered as an explicit conditioning variable through standardized log(1 + Δ*t*). Subject-wise train/validation/test splitting prevented leakage between pairs from the same individual. The model used 1,916 consecutive pairs from 491 subjects; all four latent dimensions were active, with an effective latent dimension of 3.93. On held-out test pairs, median reconstruction MSE was 0.00808 mV^2^ and median relative MSE was 0.728. Physiological interpretation combined real feature changes between the two observed ECGs with decoder traversals reconstructed on top of real HCM reference beats. Four recurrent change modes emerged (Extended Data Table 4). *D*_*z*00_ represented precordial/lateral repolarization change and was most strongly associated with change in V4 T-wave amplitude (*ρ* = −0.510, global FDR *q* = 1.81 × 10^*−*17^; traversal *R*^2^ = 0.981). *D*_*z*01_ represented coupled precordial depolarization–ST/T change and was anchored by V3 net QRS-area change (*ρ* = −0.694, *q* = 3.27 × 10^*−*38^; traversal *R*^2^ = 0.894). *D*_*z*02_ represented lateral/LV-oriented depolarization-force change and was associated with V5 net QRS-area change (*ρ* = 0.381, *q* = 1.39 × 10^*−*9^; traversal *R*^2^ = 0.991). *D*_*z*03_ represented precordial activation redistribution/R–S balance change and was anchored by V3 net QRS-area change (*ρ* = −0.529, *q* = 4.95 × 10^*−*19^; traversal *R*^2^ = 0.872). Their selected physiological anchor associations showed consistent direction across training, validation and test splits and were concordant with decoder traversals (Fig. 5).

**Figure 5:**
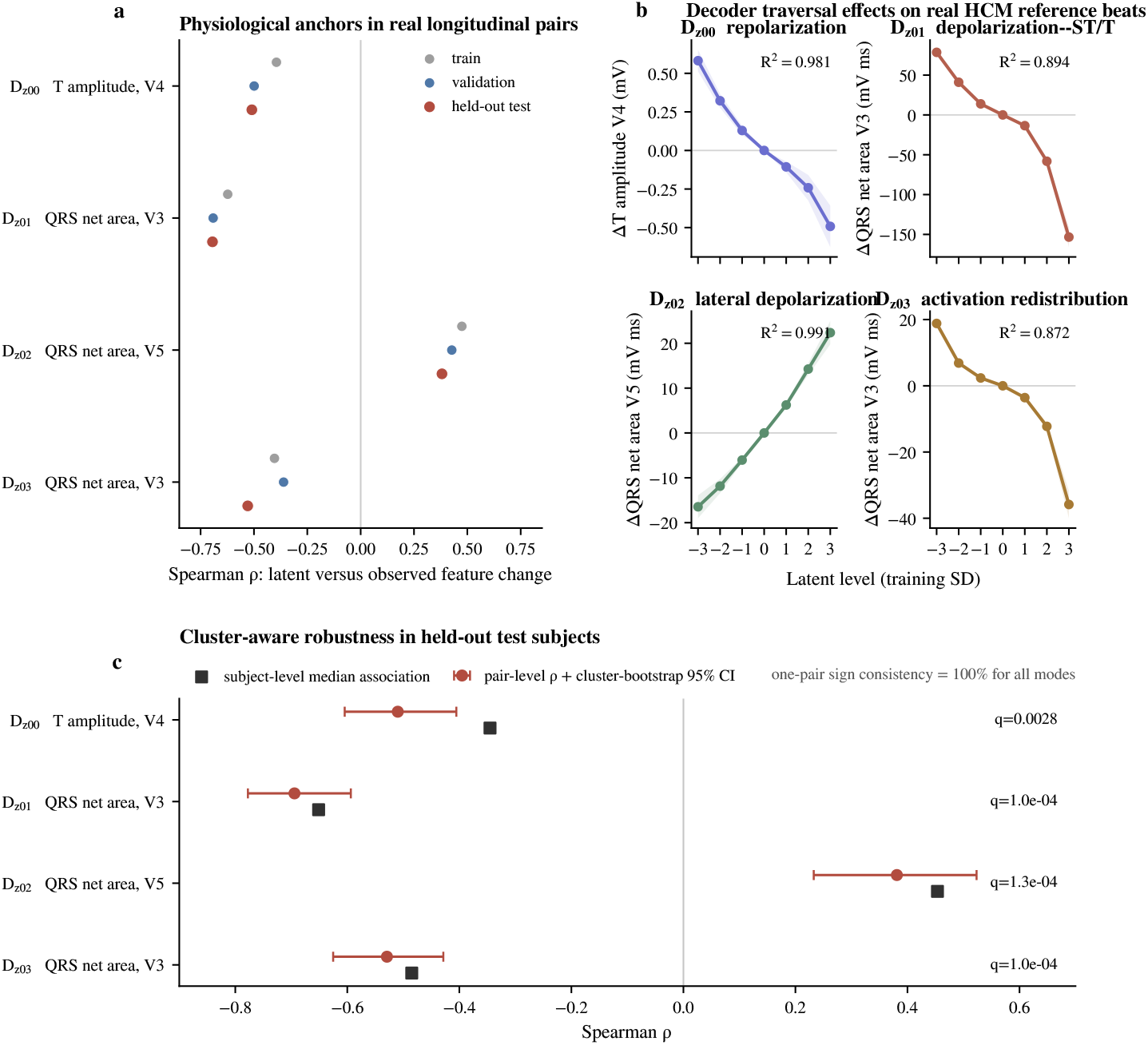
The longitudinal Delta-VAE identifies four interpretable and robust modes of within-subject ECG change. **a**, Associations between the four longitudinal latent coordinates and their selected physiological ECG-feature changes across training, validation and held-out test pairs. **b**, Decoder traversal effects for the four physiological anchors, obtained by varying each latent coordinate while holding the remaining coordinates and inter-visit interval fixed and measuring feature changes after adding the decoded waveform change to real HCM reference beats. **c**, Cluster-aware assessment of the four selected anchor associations in held-out test subjects, showing pair-level Spearman correlations with subject-cluster bootstrap 95% confidence intervals and subject-level median associations.

Because multiple consecutive pairs can arise from one subject, the four selected anchor associations were additionally assessed using subject-cluster bootstrap, subject-level median associations with permutation testing, and repeated one-pair-per-subject resampling. All four retained 95% cluster-bootstrap confidence intervals excluding zero, FDR-adjusted subject-level permutation *q <* 0.05, and 100% one-pair-per-subject sign consistency. These analyses are cluster-aware robustness analyses of selected physiological anchors rather than independent external validation. As a sensitivity analysis, a separately trained interval-normalized rate VAE recovered related depolarization and repolarization change domains despite the expected permutation, inversion or rotation of independently learned latent axes. Reconstruction of held-out waveform change was less efficient than with the primary time-conditioned Delta-VAE (median relative MSE 0.857 versus 0.728), supporting the use of the signed, non-rate-normalized waveform difference Δ*X* with explicit Δ*t* conditioning as the primary longitudinal formulation.

### 3.7. Change modes align with patient-specific longer-term manifold trajectories

We next integrated the pairwise Delta-VAE representation with patient-specific slopes from the frozen-manifold mixed-effects analysis. For each subject, the signed median of each Delta-VAE latent coordinate across consecutive pairs was compared with patient-specific longitudinal slopes estimated as best linear unbiased predictors (BLUPs) for 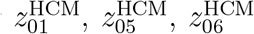 and *d*_*N*_ . Six of 16 tested associations survived FDR correction across 491 overlapping subjects. The strongest association linked the repolarization-change mode *D*_*z*00_ to the 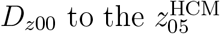 trajectory (*ρ* = 0.330, FDR *q* = 9.68 × 10^*−*13^). The lateral depolarization-force mode *D*_*z*02_ was also aligned with 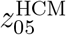 (*ρ* = 0.235, FDR *q* = 1.13 × 10^*−*6^). The coupled precordial depolarization–ST/T mode *D*_*z*01_ was negatively associated with the 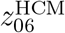 trajectory, whereas the activation-redistribution mode *D*_*z*03_ was associated with both the 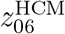 trajectory and increasing distance from the Normal manifold (*d*_*N*_ : *ρ* = 0.162, FDR *q* = 0.00166). All six primary associations preserved their direction in the model-independent OLS sensitivity analysis among 376 subjects with at least three visits, in which six of 16 associations were again FDR significant.

The held-out Delta-VAE test subset contained 73 subjects and did not yield FDR-significant associations after correction across 16 tests, although the dominant effects preserved similar directions and magnitudes. For example, the 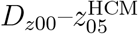 association was *ρ* = 0.324 in held-out subjects. We therefore interpret this analysis as evidence of alignment between complementary longitudinal representations, not as statistically independent validation because both summaries derive from overlapping longitudinal ECG information (Fig. 6).

### 3.8. Sarcomeric genotype is associated with electrical phenotype position but not with detectable trajectory-rate differences

A prespecified genetic analysis classified subjects as sarcomere-positive (Sarc+) when an exact non-ambiguous mapping identified at least one pathogenic or likely pathogenic variant in a core sarcomeric gene, and as sarcomerenegative (Sarc−) when an exact non-ambiguous mapping had a negative genetic result and no pathogenic or likely pathogenic variant. The primary longitudinal genetic cohort comprised 320 subjects (195 Sarc+ and 125 Sarc−) and 1,720 visits. Mixed-effects models tested genotype-by-time interactions for 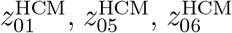 and *d*_*N*_, with adjustment for baseline age and a baseline-age-by-time interaction. No genotype-by-time interaction remained significant after FDR correction across the four selected outcomes. In secondary analyses of the baseline genotype effects from the same age-adjusted longitudinal mixed-effects models, evaluated at the cohort mean baseline age, sarcomeric genotype was associated with position along selected HCM-oriented electrical phenotype coordinates. Sarc+ subjects had lower baseline 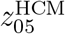values than Sarc− subjects (Sarc+ minus Sarc− effect = −0.654, 95% CI −0.902 to −0.407, nominal *p* = 2.2 × 10^*−*7^) and nominally higher base-line 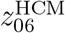 values (effect = 0.310, 95% CI 0.054–0.566, nominal *p* = 0.0177). These findings suggest that sarcomeric genotype is more clearly related to electrical phenotype position than to the average rate of longitudinal movement through the selected ECG representations. This result does not imply absence of genotype-dependent biological progression.

### 3.9. External evaluation in MIMIC-IV replicates HCM-manifold transport and longitudinal 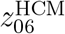 dynamics

We next evaluated the frozen SHaRE representations in the independent HCM cohort derived from MIMIC-IV (Extended Data Table 4). At baseline, 98.2% of MIMIC subjects fell within the empirical 95% coverage boundary of the HCM training distribution, closely matching the 96.5% observed in SHaRe, whereas 54.6% of MIMIC subjects and 49.7% of SHaRe subjects lay outside the corresponding Normal-reference boundary. The prespecified 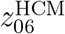 phenotype was similarly distributed across cohorts (SHaRe mean±SD, 0.457 ± 1.147; MIMIC, 0.358 ± 1.107), with a standardized external-minus-internal mean shift of −0.086 SD (95% bootstrap CI −0.224 to 0.042; FDR-adjusted KS *q* = 0.736). These findings indicate that the HCM-oriented latent representation, and particularly 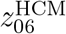, transported to the external cohort without refitting of the model or latent reference distributions. Longitudinal replication was assessed after consolidating temporally clustered MIMIC ECGs into bounded 30-day clinical windows. Among 258 subjects contributing 2,486 representative visits, 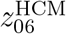 decreased by 0.0466 latent units per year (95% CI −0.0670 to −0.0263, *p* = 7.1 × 10^*−*6^), closely matching the SHaRe estimate of −0.0531 units per year. The corresponding standardized slopes were −0.0422 and −0.0456 baseline SD units per year in MIMIC and SHaRe, respectively. The estimated external-minus-internal slope difference was 0.0065 units per year (95% CI −0.0183 to 0.0313), although this comparison was not interpreted as a formal equivalence test. The external slope remained stable across alternative 7- and 90-day visit windows, restriction to subjects with at least three visits or at least one year of follow-up, and an analysis retaining all available ECGs. A subject-balanced sensitivity analysis further showed a median individual 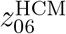 slope of −0.0360 units per year (95% subject-bootstrap CI −0.0634 to −0.0167), with 61.6% of subjects exhibiting negative slopes (two-sided exact sign-test *p* = 2.26 × 10^*−*4^). First-to-last annualized changes yielded a similar median slope of −0.0397 units per year (95% bootstrap CI −0.0629 to −0.0142). Thus, the longitudinal evolution of 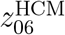 was independently reproduced in MIMIC-IV and was not attributable to unequal ECG sampling density across subjects.

## 4. Discussion and conclusions

We developed an ECG-first latent-manifold framework that separates deviation from physiological ECG morphology, heterogeneity within established HCM and within-subject electrical change over time. HCM ECGs overlapped the physiological reference manifold while extending towards its periphery (see Fig. 2), whereas the independently learned HCM-oriented manifold organized this heterogeneity along interpretable morphological axes that largely recombined physiological modes (see Fig. 3; Extended Data Table 4). Serial ECGs revealed distinct population-level trajectories, with 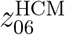 showing the strongest and most robust longitudinal change, including after censoring major clinical transitions (see Fig. 4; Table 1). Lower 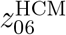 was also associated with subsequent heart-failure risk (Table 2), while a complementary Delta-VAE identified recurrent modes of within-subject waveform change that aligned with longer-term manifold trajectories (see Figs. 5 and 6; Extended Data Table 4). Finally, the HCM-oriented representation and the negative longitudinal 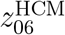 trajectory were reproduced in an independent MIMIC-IV cohort (see Extended Data Table 4). Because all latent representations were learned without supervision from genotype, clinical severity or outcomes, these findings support their interpretation as electrical phenotypes rather than proxies explicitly optimized for external clinical targets.

**Figure 6:**
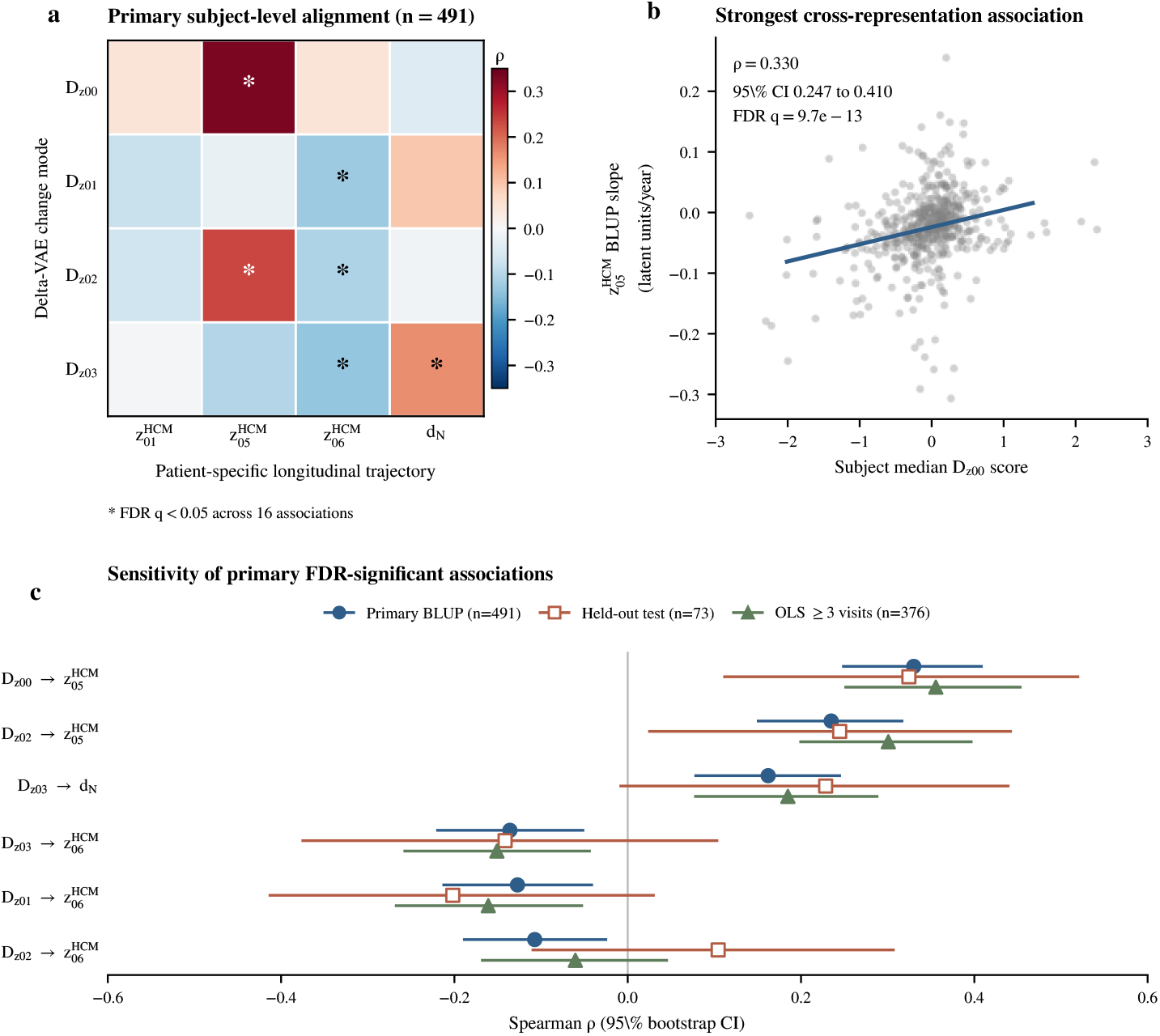
Within-subject ECG change modes align with longer-term patient-specific electrical trajectories. **a**, Subject-level Spearman associations between median signed Delta-VAE coordinates and patient-specific longitudinal slopes in the frozen HCM-oriented manifold and distance from the Normal manifold. Asterisks indicate associations surviving FDR correction across the 16 prespecified comparisons. **b**, Strongest cross-representation association, linking the repolarization-change mode *D*_*z*00_ with the patient-specific slope of 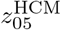. The fitted line is shown for visualization only; statistical inference is based on the Spearman association across all subjects. Extremely isolated BLUP slopes were omitted from visualization only and retained in the statistical analysis. **c**, Comparison of the six primary FDR-significant associations across the primary BLUP analysis, held- out Delta-VAE test subjects and the model-independent empirical OLS sensitivity analysis restricted to subjects with at least three visits. Points denote Spearman correlations and horizontal lines indicate bootstrap 95% confidence intervals. These analyses quantify alignment between complementary longitudinal representations and are not interpreted as independent validation because both representations derive from overlapping longitudinal ECG information.

Previous computational work has already established that HCM contains heterogeneous electrical phenotypes. Computational QRS and T-wave biomarkers combined with unsupervised clustering have been used to identify discrete HCM ECG subgroups and to relate these groups to patterns of hypertrophy and arrhythmic-risk markers [16]. More recently, supervised deep-learning approaches have shown that 12-lead ECG morphology can also distinguish predefined anatomical HCM phenotypes, including septal and apical hypertrophic patterns [28]. Our findings extend these approaches in two directions. First, rather than optimizing ECG representations for pre defined anatomical classes or partitioning patients into discrete phenotypic groups, the HCM-oriented manifold represents variation continuously along interpretable morphological axes learned directly from the multilead wave-form. Second, this continuous representation provides a natural coordinate system for longitudinal analysis, allowing serial ECGs from the same patient to be represented as trajectories through electrical phenotype space rather than as transitions between predefined classes or clusters.

The cross-manifold analysis further places HCM electrical heterogeneity in relation to physiological variation. Latent representations are increasingly being used as quantitative biomedical phenotypes [13]. In ECG analysis, latent-space decoding has been used to interpret features associated with predefined high-risk HCM phenotypes [3], while VAE-derived embeddings have been incorporated into supervised models for left ventricular hypertrophy classification [10]. Our framework instead treats latent representations as unsupervised electrical phenotype spaces and directly compares an HCM-oriented geometry with a separately learned physiological reference. The high cross-manifold predictability therefore suggests that much of HCM electrical heterogeneity reflects reorganization of physiological modes rather than a wholly distinct disease-specific latent space.

Previous AI-ECG studies have shown that supervised HCM scores can track treatment response and longitudinal changes in clinically defined disease features. HCM probability scores decreased during mavacamten treatment and correlated with haemodynamic and biomarker changes [25], while probabilities trained to detect high-risk echocardiographic features followed changes in structural-risk status over time [19]. Our framework instead represents longitudinal electrical evolution without defining the expected clinical target in advance: frozen-manifold trajectories describe longer-term movement through phenotype space, whereas the Delta-VAE identifies recurrent modes of waveform change between consecutive visits. This distinction is particularly evident in the longitudinal HCM-oriented coordinates. The three population-level trajectories were not interchangeable, and the event-censoring analyses indicate that they have different sensitivities to clinically important transitions. Longitudinal change along the lateral/frontal QRS-force coordinate 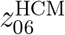 remained significant after censoring septal reduction therapy, atrial fibrillation and pacemaker implantation, including when these transitions were considered jointly. By contrast, changes in 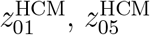 and particularly distance from the Normal manifold were attenuated after censoring septal reduction therapy. This observation is biologically plausible because septal reduction interventions themselves can substantially modify ventricular activation: both myectomy and alcohol septal ablation are known to produce major QRS and conduction changes, including frequent new bundle-branch block [23]. Event censoring cannot distinguish intervention effects from the disease state that led to intervention and also changes the amount and composition of follow-up. These analyses should therefore not be interpreted causally. Instead, they indicate that different electrical coordinates capture distinct components of longitudinal evolution, arguing against reduction of serial ECG change to a single global progression score.

Prognostic information from the conventional ECG in HCM has been recognized previously, with specific QRS and repolarization abnormalities associated with adverse cardiovascular outcomes [2]. Our findings extend this concept to a continuous multilead electrical phenotype learned without outcome supervision and identified initially through its longitudinal behaviour. Lower 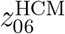 was associated with subsequent heart-failure risk both at baseline and as a time-updated exposure, suggesting that clinically relevant information can emerge from the unsupervised organization of ECG morphology rather than from predefined ECG abnormalities alone. Thus, the lateral/frontal QRS-force phenotype represented by 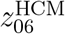 is not only longitudinally robust but also carries information about subsequent heart-failure risk. Importantly, the consistency between baseline and time-updated analyses indicates that serially available 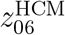 values retain risk information, but does not establish that within-patient decline in 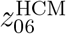 itself causes or independently predicts increasing risk. The association with ventricular-arrhythmic events was directionally consistent but should remain exploratory because of the limited number of events. The available data did not establish residual post-treatment risk. Landmark analyses after septal reduction therapy contained too few subsequent events for reliable inference, despite substantial peri-procedural variation in 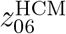. Accordingly, the present results support 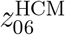 as a risk-associated electrical phenotype, particularly for heart failure, rather than as a validated post-treatment risk marker. Larger cohorts with denser post-intervention ECG follow-up will be required to determine whether residual or evolving 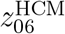provides prognostic information after septal reduction therapy.

The Delta-VAE provides a complementary description of this evolution at shorter temporal scale. Its four modes captured recurrent changes in repolarization, coupled depolarization–ST/T morphology, lateral depolarization forces and precordial activation redistribution. Physiological associations derived from real ECG pairs were concordant with decoder traversals and remained supported when repeated observations within subjects were explicitly accounted for. Related depolarization and repolarization domains were also recovered by an independently initialized interval-normalized rate VAE, despite the expected permutation, inversion or rotation of independently learned latent axes. The less efficient reconstruction of the rate-normalized model supports the primary formulation in which the signed waveform difference Δ*X* is represented directly while elapsed time is supplied separately as a conditioning variable. Together with the alignment between Delta-VAE summaries and patient-specific manifold slopes, these results suggest that local waveform change and longer-term electrical drift are different temporal views of an underlying evolving phenotype rather than unrelated representations.

The genetic findings should also be interpreted in the context of established HCM biology. Electrical abnormalities can precede overt hypertrophy in sarcomere-mutation carriers [15], and ECG-based deep-learning models have shown that sarcomeric genotype can be partially inferred from electrical morphology [4]. Large SHaRe analyses have further shown that pathogenic sarcomeric variants are associated with earlier disease expression and substantially greater lifetime clinical burden [11]. Against this background, our finding that sarcomeric genotype was associated with selected baseline electrical coordinates but did not significantly modify the average longitudinal slopes is not evidence that genotype is unrelated to disease progression. Rather, it suggests that genotype-associated differences in electrical phenotype position and the subsequent rate of movement through the ECG manifold are separable quantities. Genetic substrate may influence when and where an electrical phenotype becomes expressed without necessarily producing a detectable uniform difference in annual ECG trajectory over the follow-up interval and sample size available here.

External validation provides a complementary form of evidence. Supervised AI-ECG models for HCM detection have shown substantial generalizability across independent international cohorts [26], including external validation in MIMIC-IV [24]. Here, external evaluation addressed a different question: whether a frozen unsupervised HCM phenotype representation remains applicable across cohorts and whether its longitudinal behaviour is reproducible. The preserved manifold coverage and independent replication of the negative 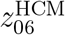 trajectory in MIMIC-IV support transportability of both phenotype position and temporal evolution.

This study has limitations. The Normal and HCM cohorts originated from different source datasets, so between-source reconstruction differences and descriptive separation metrics cannot be interpreted as diagnostic performance. External evaluation in MIMIC-IV supported transport of the frozen HCM-oriented representation and independently replicated the longitudinal 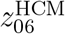 trajectory, but did not extend to all components of the framework. In particular, the clinical-risk association of 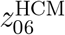, genotype relationships and the Delta-VAE change modes were not externally validated. The Delta-VAE represents unsupervised waveform change rather than a clinically validated progression score. Patient-specific BLUP slopes and Delta-VAE summaries are derived from overlapping serial ECG observations; their association therefore demonstrates alignment between complementary representations rather than statistically independent validation. The clinical-risk analyses of 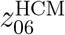 were secondary and were adjusted for age and sex but not for the full set of established structural, haemodynamic and clinical risk markers; they therefore demonstrate risk association rather than independent or incremental predictive value. Similarly, the time-updated analyses show that serially available 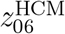 values retain information about subsequent risk, but do not establish that within-patient decline in 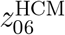 itself causes or independently predicts increasing risk. Ventricular-arrhythmic analyses were limited by the small number of events, and post-SRT landmark analyses were underpowered. Event-censoring analyses reduce available follow-up and alter cohort composition and cannot distinguish effects of clinical interventions from the disease states that prompted those interventions. Finally, the median-beat representation intentionally emphasizes morphology and does not represent rhythm or beat-to-beat variability as primary latent information.

Together, these results position the serial ECG as more than a binary marker of HCM presence. Established electrical abnormalities can be organized within a continuous geometry referenced to physiological variation, and individual patients can be followed as they move through this electrical phenotype space. Within this representation, *z*^HCM^ emerged as a longitudinally robust electrical phenotype whose lower values were associated with subsequent heart-failure risk, providing an initial link between unsupervised electrical evolution and clinical outcome. By separating physiological reference position, within-HCM phenotype organization, within-subject change and subsequent clinical association, the framework provides a state-and-change representation that complements supervised AI-ECG detection and monitoring approaches. Future work should determine the structural substrates of these electrical trajectories and whether they provide incremental prognostic information beyond established imaging, haemodynamic, genetic and clinical risk markers.

## Ethics and consent

The SHaRe registry was conducted under the applicable institutional review board and ethics approvals, with de-identified clinical data collected according to the consent or waiver requirements of the participating institutions. The present study analysed de-identified SHaRe data made available to Politecnico di Milano within the EU-funded SMASH-HCM project, in accordance with the applicable data-use and governance requirements.

## Data availability

The PTB-XL data used as the Normal-reference cohort are publicly available from the original data repository. The HCM clinical and ECG data used in this study derive from the SHaRe registry and are subject to the corresponding data-governance, consent and data-use requirements; these individual-level data therefore cannot be redistributed by the authors. Access to SHaRe data is subject to approval by the relevant data-governance bodies and applicable data-use agreements. Derived non-identifying analysis outputs required to reproduce the reported results are provided, where permitted, through the study repository. The dataset is not publicly available because it contains electronic health records. Sharing these data externally without proper consent could compromise patient privacy and violate the institutional review board’s approval for the study. However, deidentified data may be made available upon reasonable request to the corresponding author, subject to institutional and ethical approvals by the review board.

## Code availability

The analysis code used to generate the results reported in this study is publicly available through the study repository at: https://github.com/Hal9000-lab/SMASH-HCM-ECG.

The repository includes the frozen analysis code, model configurations, deterministic data-split definitions, environment specifications and scripts required to reproduce the reported figures and tables from authorized input data. Source clinical data subject to SHaRe governance restrictions are not redistributed through the repository.

## Acknowledgments

N/A

## Author contributions

P.C.: Conceptualization, Methodology, Data Analysis, Supervision, Writing – original draft, Writing – review and editing. M.T.: Data Analysis, writing – review and editing. V.C. Data Analysis, Writing – review and editing. L.M.: Conceptualization, funding acquisition, project administration, Writing – review and editing. All authors reviewed and approved the final manuscript.

## Supplementary Materials

**Extended Data Fig. 1.**
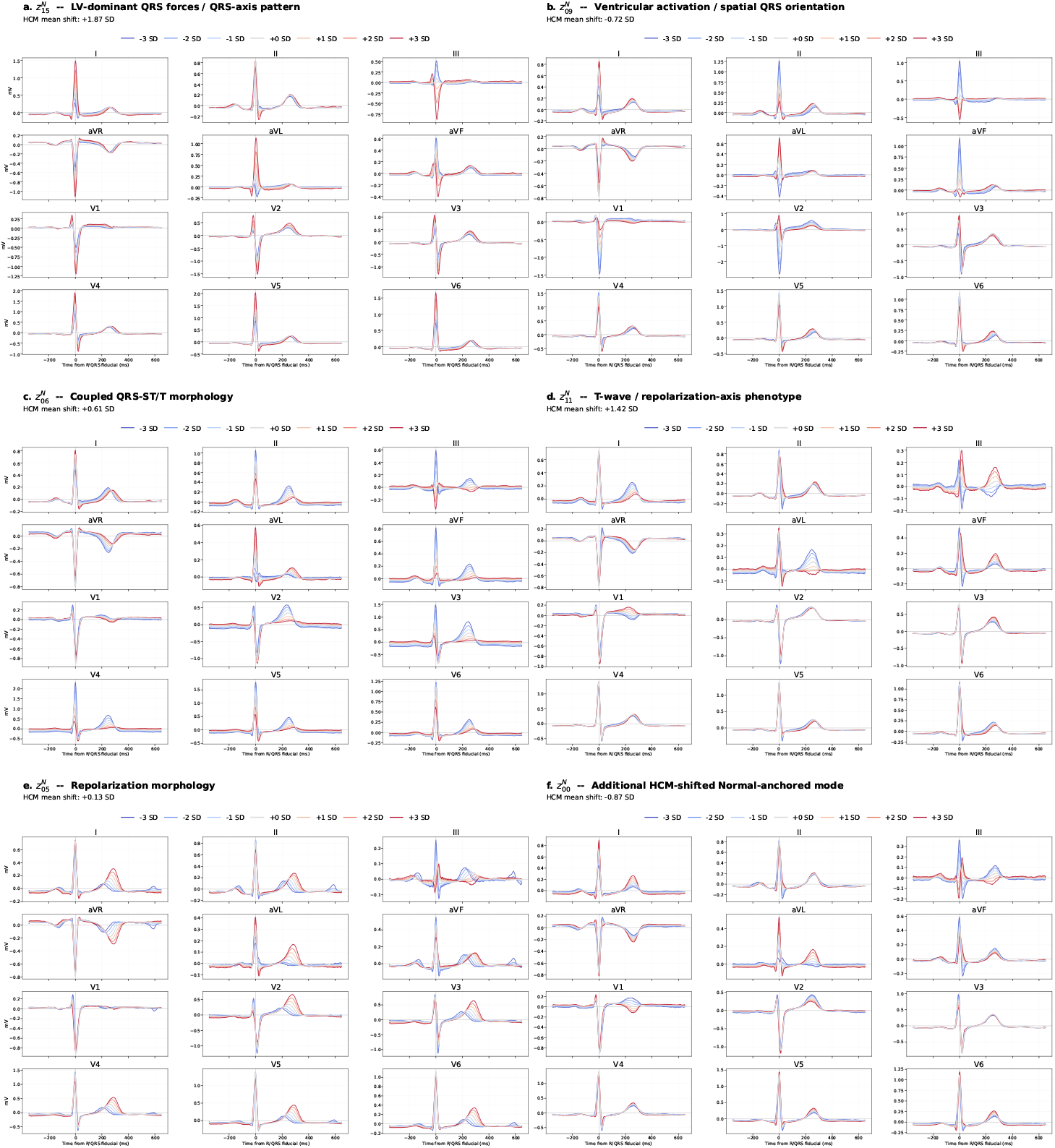
Principal Normal-anchored ECG deformation modes associated with HCM. Decoder traversals show six Normal-anchored latent coordinates prominently displaced in HCM. For each coordinate, *z*^*N*^ was varied from −3 to +3 empirical Normal-training SD around the Normal-training latent centroid, while all remaining latent coordinates were fixed at the centroid. The corresponding 12-lead median beat was decoded at each traversal level. Values above each panel indicate the mean HCM displacement along that coordinate, expressed in Normal-training SD units.

**Extended Data Fig. 2.**
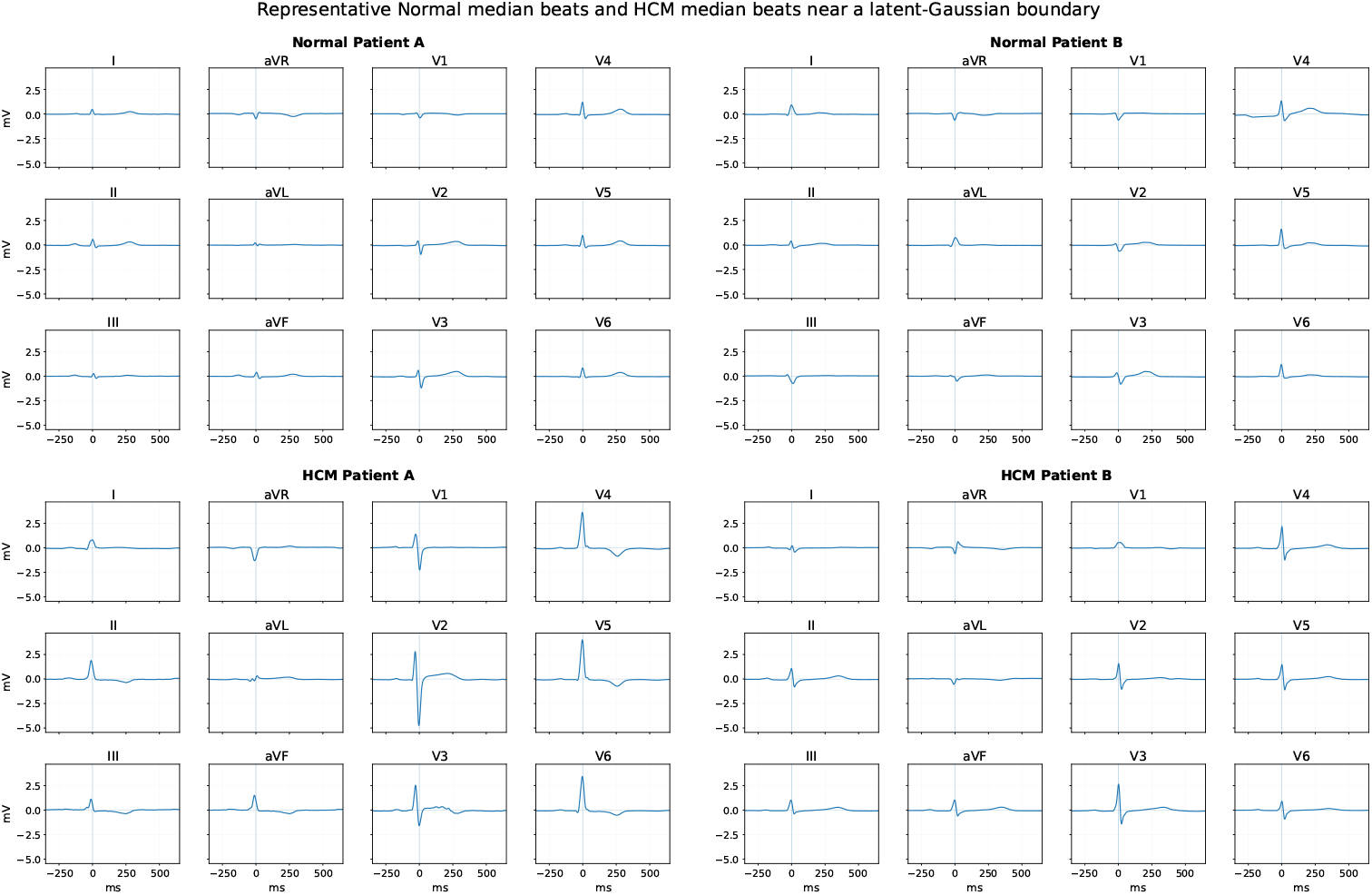
Representative 12-lead median-beat ECG morphologies in Normal-reference and HCM subjects. Four representative subjects are shown in a 2 × 2 arrangement, with two Normal-reference subjects on the top and two HCM subjects on the bottom. Normal subjects were selected close to the central region of the Normal-anchored latent distribution, whereas HCM subjects were selected near the 95% boundary of the HCM-oriented latent distribution to illustrate peripheral, but non-extreme, HCM phenotypes. Each panel displays the observed median beat for all 12 ECG leads using the original physical amplitudes in mV, without waveform reconstruction or amplitude normalization. Common axis scaling facilitates direct comparison of voltage and morphology across subjects.

**Extended Data Table 1.** Baseline clinical characteristics of the exact ECG-to-SHaRe mapped HCM cohort. Continuous variables are reported as median [interquartile range] and categorical variables as *n* (%). Baseline was defined by the first valid waveform ECG. Echocardiographic variables were taken from the nearest examination within 7 days of the baseline ECG. Percentages are calculated among subjects with available data. SRT denotes septal reduction therapy.

| Characteristic | Available *n* | Value |
|---|---|---|
| Subjects | 623 | 623 |
| Age at baseline ECG, years | 623 | 50.7 [37.9–61.3] |
| Female sex | 623 | 226 (36.3%) |
| Valid ECGs per subject | 623 | 3 [2–5] |
| Waveform follow-up, years | 623 | 2.39 [0.53–6.82] |
| Clinical follow-up, years | 619 | 8.09 [6.02–12.97] |
| *Echocardiography within 7 days of baseline ECG* |  |  |
| Left ventricular ejection fraction, % | 502 | 66 [62–70] |
| Maximum LV wall thickness, mm | 556 | 17 [15–21] |
| Left atrial volume index, ml m<sup>−2</sup> | 505 | 41.1 [32.7–52.3] |
| LVOT gradient ≥ 50 mmHg at rest or provocation | 516 | 88 (17.1%) |
| *Clinical history at or before baseline ECG* |  |  |
| Atrial fibrillation | 619 | 60 (9.7%) |
| Implantable cardioverter-defibrillator | 622 | 34 (5.5%) |
| Septal reduction therapy | 623 | 21 (3.4%) |
| Heart-failure composite | 623 | 34 (5.5%) |
| *Genetic classification* |  |  |
| Classifiable for the primary Sarc+ versus Sarc− definition | 623 | 386 (62.0%) |
| Sarc+, among classifiable subjects | 386 | 240 (62.2%) |
| Sarc−, among classifiable subjects | 386 | 146 (37.8%) |

**Extended Data Table 2.** Physiological interpretation of the HCM-oriented latent coordinates. For each latent coordinate, a representative ECG feature is reported together with its Spearman association with the latent coordinate in HCM training ECGs and the corresponding decoder-traversal effect. Traversals varied one latent coordinate while holding the remaining coordinates fixed and were evaluated across −3 to +3 empirical training-set SD. Concordant associations between real ECGs and decoder traversals were used to support physiological interpretation.

| Latent | Interpretation | Representative ECG feature | Lead | Spearman *ρ* | Traversal slope/SD | Traversal *R*<sup>2</sup> |
|---|---|---|---|---|---|---|
| *z*<sub>00</sub><sup>HCM</sup> | Precordial QRS-force redistribution | QRS net area | V2 | −0.484 | −14.19 | 0.972 |
| *z*<sub>01</sub><sup>HCM</sup> | Precordial R/S transition balance | R/S balance | V4 | +0.329 | +0.226 | 0.982 |
| *z*<sub>02</sub><sup>HCM</sup> | Precordial repolarization | T-wave amplitude | V4 | +0.732 | +0.324 | 0.998 |
| *z*<sub>03</sub><sup>HCM</sup> | Precordial depolarization force | QRS net area | V4 | +0.687 | +15.27 | 0.998 |
| *z*<sub>04</sub><sup>HCM</sup> | Global QRS magnitude | Global QRS peak-to-peak amplitude | Global | +0.685 | +0.728 | 0.926 |
| *z*<sub>05</sub><sup>HCM</sup> | Lateral/LV-oriented depolarization force | QRS net area | V5 | +0.817 | +21.79 | 0.996 |
| *z*<sub>06</sub><sup>HCM</sup> | Lateral/frontal QRS-force redistribution | QRS net area | aVL | −0.663 | −20.05 | 0.887 |
| *z*<sub>07</sub><sup>HCM</sup> | Mixed precordial repolarization–activation | T-wave area | V3 | +0.512 | +26.29 | 1.000 |

**Extended Data Table 3.** Principal longitudinal Delta-VAE modes and physiological interpretations.

| Latent | Interpretation | Selected physiological anchor |
|---|---|---|
| *D*<sub>*z*00</sub> | Precordial/lateral repolarization change | ΔT amplitude, V4 |
| *D*<sub>*z*01</sub> | Precordial depolarization–ST/T coupled change | ΔQRS net area, V3 |
| *D*<sub>*z*02</sub> | Lateral/LV-oriented depolarization-force change | ΔQRS net area, V5 |
| *D*<sub>*z*03</sub> | Precordial activation redistribution/R–S balance | ΔQRS net area, V3 |

**Extended Data Table 4.**
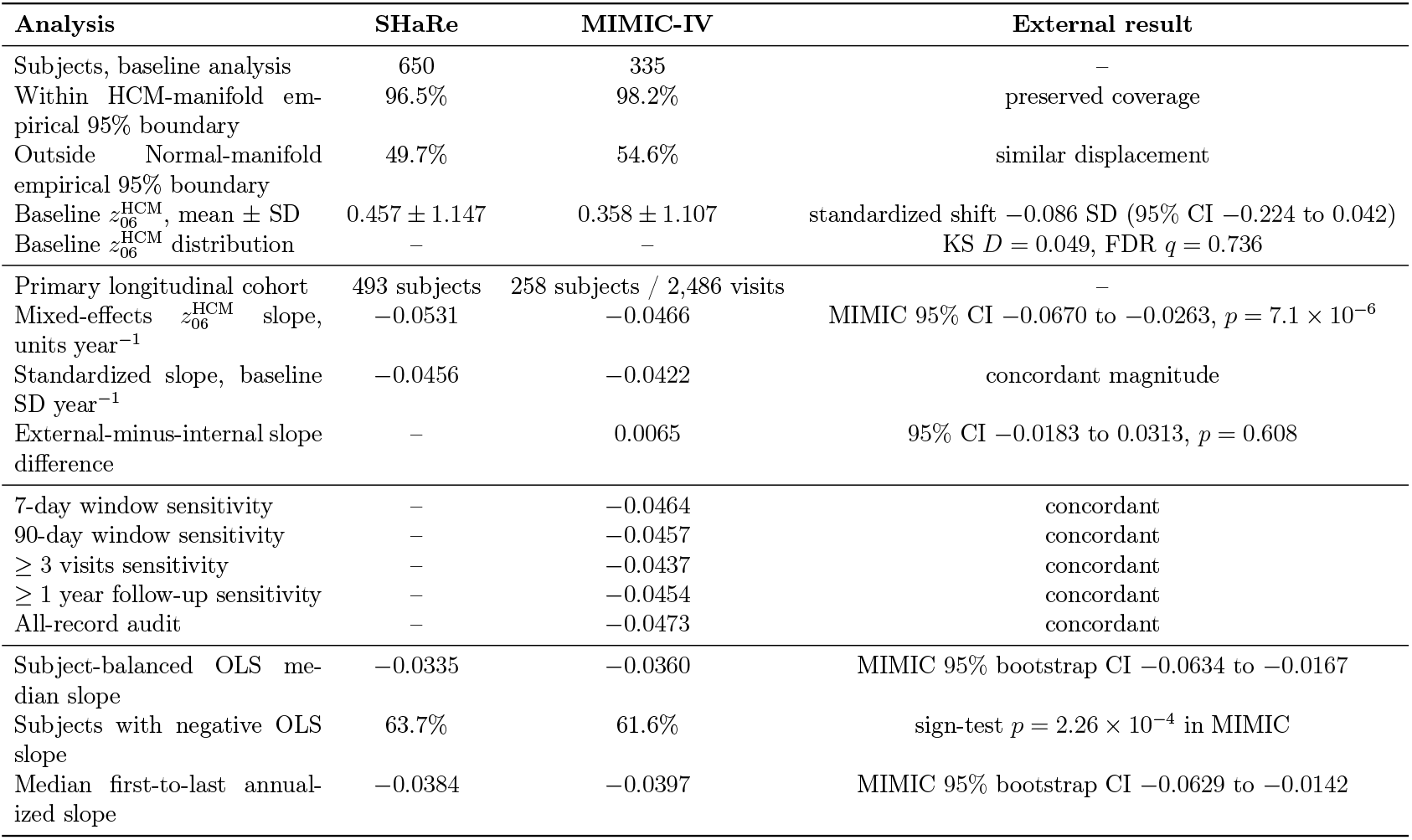
Baseline manifold coverage was defined using empirical distance thresholds estimated exclusively from the original Normal and HCM training distributions. Longitudinal MIMIC analyses used bounded 30-day clinical windows unless otherwise specified. SHaRe–MIMIC comparisons are descriptive and were not interpreted as formal equivalence tests.

| Analysis | SHaRe | MIMIC-IV | External result |
|---|---|---|---|
| Subjects, baseline analysis | 650 | 335 | – |
| Within HCM-manifold em-pirical 95% boundary | 96.5% | 98.2% | preserved coverage |
| Outside Normal-manifold empirical 95% boundary | 49.7% | 54.6% | similar displacement |
| Baseline z<sub>06</sub><sup>HCM</sup>, mean ± SD | 0.457 ± 1.147 | 0.358 ± 1.107 | standardized shift −0.086 SD (95% CI −0.224 to 0.042) |
| Baseline z<sub>06</sub><sup>HCM</sup> distribution | – | – | KS D = 0.049, FDR q = 0.736 |
| Primary longitudinal cohort | 493 subjects | 258 subjects / 2,486 visits | – |
| Mixed-effects z<sub>06</sub><sup>HCM</sup> slope, units year<sup>−1</sup> | −0.0531 | −0.0466 | MIMIC 95% CI −0.0670 to −0.0263, p = 7.1 × 10<sup>−6</sup> |
| Standardized slope, baseline SD year<sup>−1</sup> | −0.0456 | −0.0422 | concordant magnitude |
| External-minus-internal slope difference | – | 0.0065 | 95% CI −0.0183 to 0.0313, p = 0.608 |
| 7-day window sensitivity | – | −0.0464 | concordant |
| 90-day window sensitivity | – | −0.0457 | concordant |
| ≥ 3 visits sensitivity | – | −0.0437 | concordant |
| ≥ 1 year follow-up sensitivity | – | −0.0454 | concordant |
| All-record audit | – | −0.0473 | concordant |
| Subject-balanced OLS me-dian slope | −0.0335 | −0.0360 | MIMIC 95% bootstrap CI −0.0634 to −0.0167 |
| Subjects with negative OLS slope | 63.7% | 61.6% | sign-test p = 2.26 × 10<sup>−4</sup> in MIMIC |
| Median first-to-last annual-ized slope | −0.0384 | −0.0397 | MIMIC 95% bootstrap CI −0.0629 to −0.0142 |

